# From meal text to carbohydrate to postprandial glucose rise: QLoRA fine-tuning of the Qwen3-4B language model and gradient-boosting prediction in type 2 diabetes

**DOI:** 10.64898/2026.10.15.26365634

**Authors:** Mohamed Nour Bou Ali, Najoua Lassoued, Bilel Ben Amor, Salem Nasr, Mohamed Amine Aouididi

## Abstract

**Background:** In people with type 2 diabetes (T2D), postprandial glucose weighs on glycaemic control, and carbohydrate counting is difficult. Patients can describe their meal but rarely quantify it.

**Objective:** To assess (1) whether a small open large language model (LLM), fine-tuned on public data, estimates the carbohydrate content of a meal described in words, and (2) what this carbohydrate adds to the prediction of the postprandial glucose rise in two public cohorts.

**Methods:** Step 1: the Qwen3-4B LLM was fine-tuned with QLoRA on Nutrition5k dishes and compared with an automatically matched USDA food composition table and with Qwen3-4B and Qwen3-8B used without training, on test dishes (internal control) and on 100 real meals of ShanghaiT2DM patients, whose reference carbohydrate was computed from the recorded quantities. Step 2: the maximum glucose rise between 0 and 120 minutes (ΔGmax) was predicted by personalised gradient boosting (M2), compared with a spline linear regression and with the Chronos-2 foundation model, using only information available at meal start and person-level cross-validation, in ShanghaiT2DM (people with T2D recruited in two Shanghai hospitals; carbohydrate estimated by the LLM) and CGMacros (free-living adults in Santa Barbara, California; carbohydrate entered in the dataset, no text). Step 3: the model was transferred between datasets. 95% confidence intervals (CI) were obtained by cluster BCa bootstrap (2,000 resamples).

**Results:** On the 100 real meals, the carbohydrate mean absolute error (MAE) of the fine-tuned model was 12.2 [9.9, 15.2] g, almost half that of the table (22.6 g; paired difference -10.5 [-16.8, -6.0] g) and nearly a third of that of the same model without fine-tuning (34.3 g), with a bias of -10.3 g; on test dishes it was 2.2 [2.0, 2.5] g. Personalised gradient boosting (M2) reached a correlation between predicted and observed rises of r = 0.63 [0.55, 0.69] in CGMacros (1,286 meals, 45 adults) and 0.49 [0.42, 0.54] in ShanghaiT2DM (2,339 meals, 99 people with T2D), about twice the linear reference (r = 0.32 and 0.23); for a rise above 50 mg/dL, its area under the ROC curve (AUROC) was 0.80 and 0.74, versus 0.64 and 0.68 with pre-meal glucose alone. Adding carbohydrate to pre-meal glucose changed r by 0.095 [0.068, 0.128] in CGMacros and by 0.004 [-0.010, 0.017] in ShanghaiT2DM; difference between datasets: 0.091 [0.060, 0.126]. Applied from one dataset to the other, the model lost correlation (Δr = -0.11 [-0.17, -0.05] towards ShanghaiT2DM and -0.12 [-0.19, -0.04] towards CGMacros). The full “text → LLM → glucose” chain was evaluated in ShanghaiT2DM only.

**Conclusion:** An open 4-billion-parameter LLM, fine-tuned with QLoRA on public data and usable offline, almost halves the error of carbohydrate counting for real meals described in words compared with an automatically matched table, with an underestimation of large meals. Personalised gradient boosting, based on pre-meal glucose and the person’s usual response, doubles the correlation of a linear reference for predicting the glucose rise in both cohorts. Carbohydrate improved this prediction in CGMacros but not in ShanghaiT2DM; the two datasets differ in many respects, and the cause of this gap is not established.

## 1. Introduction

In people with T2D, postprandial hyperglycaemia contributes to overall hyperglycaemia, and all the more as glycaemic control approaches target: in the study by Monnier et al., the share of postprandial excursions in the daily glucose excess fell from 69.7% in patients in the lowest HbA1c quintile to 30.5% in those in the highest quintile, fasting glucose taking over as control worsens [3]. Guidelines therefore make it a therapeutic target [4]. The main lever is diet, especially the amount of carbohydrate, but carbohydrate counting is difficult and imprecise, even in trained adults with type 1 or type 2 diabetes [5, 6]. Continuous glucose monitoring improves glycaemic control, including in people with T2D treated with basal insulin alone, without mealtime insulin [7], and self-management mobile applications reduce HbA1c modestly but reproducibly [8]: the missing link remains turning a meal into grams of carbohydrate. Diabetes affects about 1.4 million adults in Tunisia [9], where this work is conducted.

Two approaches have emerged. The first is personalised nutrition: the glycaemic response to the same meal varies widely between people [10] and can be predicted from the meal, glucose and individual characteristics [11–13], with a glycaemic benefit shown in a randomised trial in people with prediabetes [14] and, in a pilot trial, in people with newly diagnosed T2D [15]. The second is automatic nutrient estimation by large language models (LLMs, artificial-intelligence programs that read and write text), whose medical capabilities are progressing rapidly [16, 17]: for meals described in text, the estimates of commercial models approach those of clinicians, with differences between models [18]; they are less reliable from photographs of mixed dishes [19], and public benchmarks have been proposed [20].

Three questions remain open. (1) Can a small **open** LLM, usable offline without data transmission, be fine-tuned for this task? (2) Does it work on **real patient texts**, translated, with foods absent from its training data? (3) Does the carbohydrate it estimates **add** to the prediction of the glucose rise, beyond the glucose value measured just before the meal? A recent study on ShanghaiT2DM reported that macronutrients alone (carbohydrate, protein, fat) poorly predict the glycaemic response and that food categories (starches, fruit, vegetables, meat…) do better [21], which calls for an explicit test of the “text → nutrients → glucose” chain.

The aim of this work was threefold: (1) to fine-tune an open LLM, Qwen3-4B (Qwen3 family, Alibaba Cloud), to convert a meal described in words into nutrients, and to compare its estimates, on meals whose reference carbohydrate is known, with those of a food composition table and of LLMs used without task-specific training (“zero-shot”); (2) to measure what carbohydrate adds to the prediction of the glucose rise in two public cohorts, ShanghaiT2DM and CGMacros; (3) to evaluate the transfer of the prediction model from one dataset to the other.

## 2. Methods

### 2.1 Design, setting, period and ethics

Development and validation study of prediction models, reported according to TRIPOD+AI [1] and TRIPOD-LLM [2]. **Development** means building a model from part of the data (choice of variables, training); **validation** means measuring its performance on data that were not used to build it: here dishes and meals held out from training (step 1), people never seen by the model (step 2) and another dataset (step 3). The protocol (definitions, splits, outcomes, analyses, step-1 pass criterion) was written and the splits frozen before the LLM was trained; it was not deposited with a public time stamp. The status of each analysis (main, secondary, exploratory or sensitivity) is given in Table S1. No new data were collected: the three datasets are public, de-identified and released under open licences, and the raw files are frozen by SHA-256 hash. ShanghaiT2DM recruited its patients from September 2019 to March 2021 (Shanghai East Hospital) and then from June to November 2021 (Shanghai Fourth People’s Hospital) [22]; the recording files are dated from September 2020 to January 2022, a discrepancy not commented on by the dataset authors. This period covers the COVID-19 pandemic. CGMacros recruited from 2021 to 2024 at the Sansum Diabetes Research Institute (Santa Barbara, California) [23]; Nutrition5k was published in 2021 without a collection date [24]. Analyses were conducted in October 2026.

#### Ethics

The ShanghaiT2DM and CGMacros data were collected by their authors with the approval of their ethics committees and participants’ consent [22, 23]; Nutrition5k contains no personal data. This work is a secondary analysis of these public de-identified data, reused under their licences, with no identifying data and no contact with any person; it was therefore not submitted to an ethics committee.

#### Time of prediction

The prediction is made at the start of the studied meal (t0). The main analysis uses only information available at t0: snacks eaten after t0 are not added to the meal, treatments are counted only in the 30 minutes before t0 (-30 to 0 min), and post-meal heart rate is not used. A sensitivity analysis allowed information posterior to meal start: snacks of the following 2 hours added to the meal, treatments from -30 to +30 min and post-meal heart rate in the secondary “watch” analysis. Two eligibility criteria remain based on the post-meal period and are flagged as such: no other meal within 90 minutes and, in CGMacros, at least half a portion eaten.

### 2.2 Data sources, role of each dataset and eligibility criteria

- **Nutrition5k** [24]: cafeteria dishes described by the list of their weighed ingredients and their nutrients computed from a density table (nutrient content per gram). No person is included: the unit is the dish. In this article, “dish” refers to a Nutrition5k dish and “meal” to a patient’s meal.
- **ShanghaiT2DM** [22]: the public archive combines two sub-datasets, ShanghaiT1DM and ShanghaiT2DM. Patients were included by the dataset authors if they had diabetes according to the 1999 WHO criteria, were older than 18 years, signed informed consent and had at least 3 days of recording. Each patient wore a FreeStyle Libre H (Abbott), a continuous glucose monitoring sensor worn on the arm, providing here one value every 15 minutes; meals, weighed, were recorded in Chinese and translated into English by the dataset authors, together with treatments and clinical characteristics. The inpatient or outpatient status of each recording is not reported.
- **CGMacros** [23, 25, 26]: normoglycaemic, prediabetic or T2D adults followed for about ten days in free-living conditions, wearing simultaneously two professional continuous glucose monitoring sensors, a Dexcom G6 Pro (Dexcom; one value every 5 minutes) and a FreeStyle Libre Pro (Abbott; one value every 15 minutes), whose readings are not displayed to the wearer. The carbohydrate, protein, fat and energy of each meal were estimated by the dataset team and are entered in the dataset. People with T2D treated with an oral agent other than metformin, a GLP-1 receptor agonist or insulin were not included. The files contain no textual description of meals (only the meal type and the path of a photograph): the LLM could not be applied, and step 2 uses the carbohydrate entered in the dataset directly.
- **USDA FoodData Central, SR Legacy** [27]: food composition table of the United States Department of Agriculture, used as a comparator and to compute the reference of the real meals.

Table 1 summarises the role of each dataset.

**Table 1.**
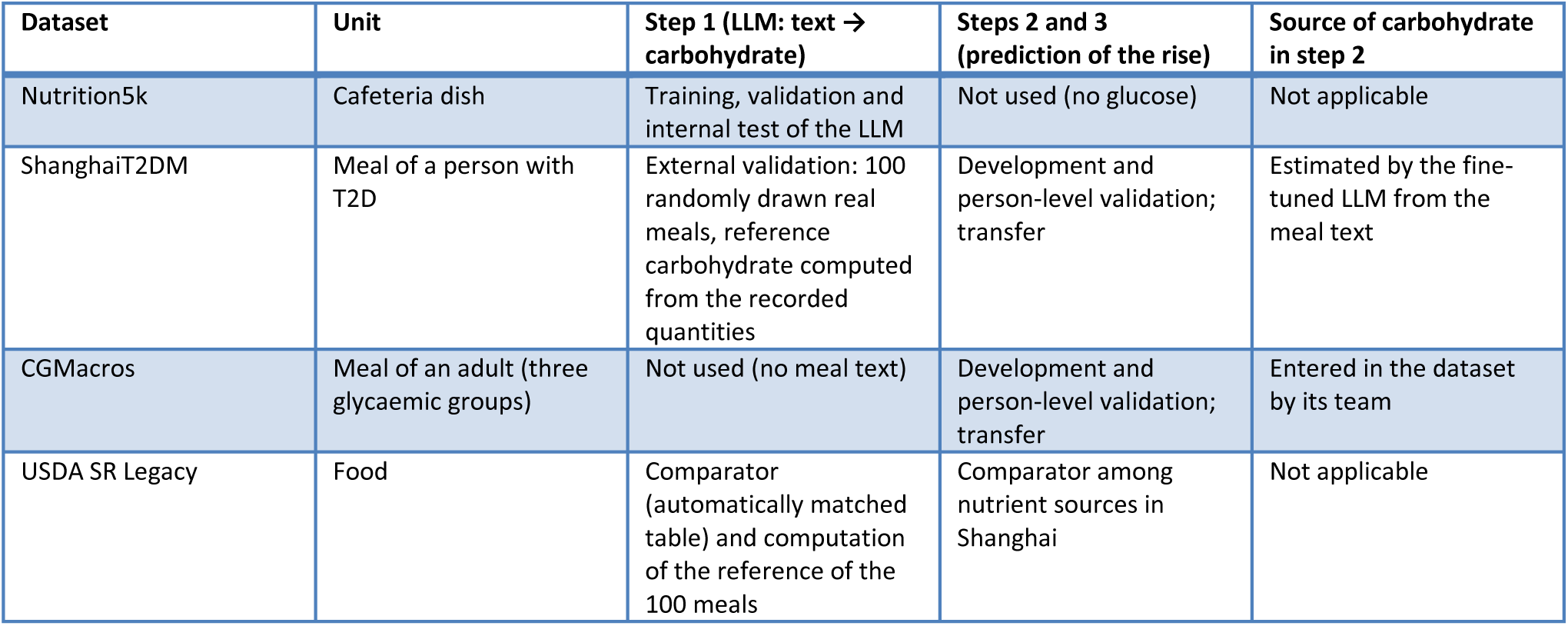
Role of each dataset in the study.

| Dataset | Unit | Step 1 (LLM: text → carbohydrate) | Steps 2 and 3 (prediction of the rise) | Source of carbohydrate in step 2 |
| --- | --- | --- | --- | --- |
| Nutrition5k | Cafeteria dish | Training, validation and internal test of the LLM | Not used (no glucose) | Not applicable |
| ShanghaiT2DM | Meal of a person with T2D | External validation: 100 randomly drawn real meals, reference carbohydrate computed from the recorded quantities | Development and person-level validation; transfer | Estimated by the fine-tuned LLM from the meal text |
| CGMacros | Meal of an adult (three glycaemic groups) | Not used (no meal text) | Development and person-level validation; transfer | Entered in the dataset by its team |
| USDA SR Legacy | Food | Comparator (automatically matched table) and computation of the reference of the 100 meals | Comparator among nutrient sources in Shanghai | Not applicable |

#### Vocabulary of meal selection

A *food intake* is any line of the dataset’s food diary, meal or snack; two lines recorded less than 15 minutes apart form a single intake. An *index meal* is an intake retained as a studied meal, as opposed to snacks. An *analysable meal* is an index meal meeting the description and glucose criteria (at least 2 glucose measurements in the 30 minutes before the meal and 7 measurements between 15 and 120 minutes after). The *annotated meals* are the 100 analysable Shanghai meals drawn at random to validate step 1.

Eligibility criteria were applied at each level, in the order of Table 2. Type 1 diabetes was excluded by design: in type 1 diabetes, the meal response depends first on the prandial insulin dose chosen by the patient, which raises a different research question. Only the ShanghaiT2DM sub-dataset was therefore extracted from the archive. The index meal was defined, in Shanghai, by the presence of a carbohydrate food found in the text by a keyword list (rice, bread, noodles…), without any LLM output, so that meal selection did not depend on the evaluated model, and, in CGMacros, which contains no text, by at least 15 g of entered carbohydrate. As each model is trained and evaluated within a single dataset, this difference does not change the rule applied to the training and testing of a given model; a sensitivity analysis also applied the same “at least 15 g” rule to both datasets (LLM carbohydrate in Shanghai).

**Table 2.**
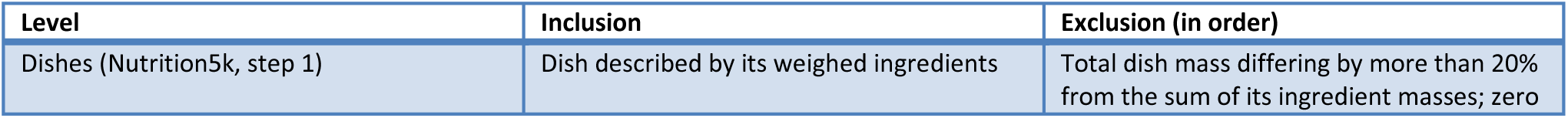
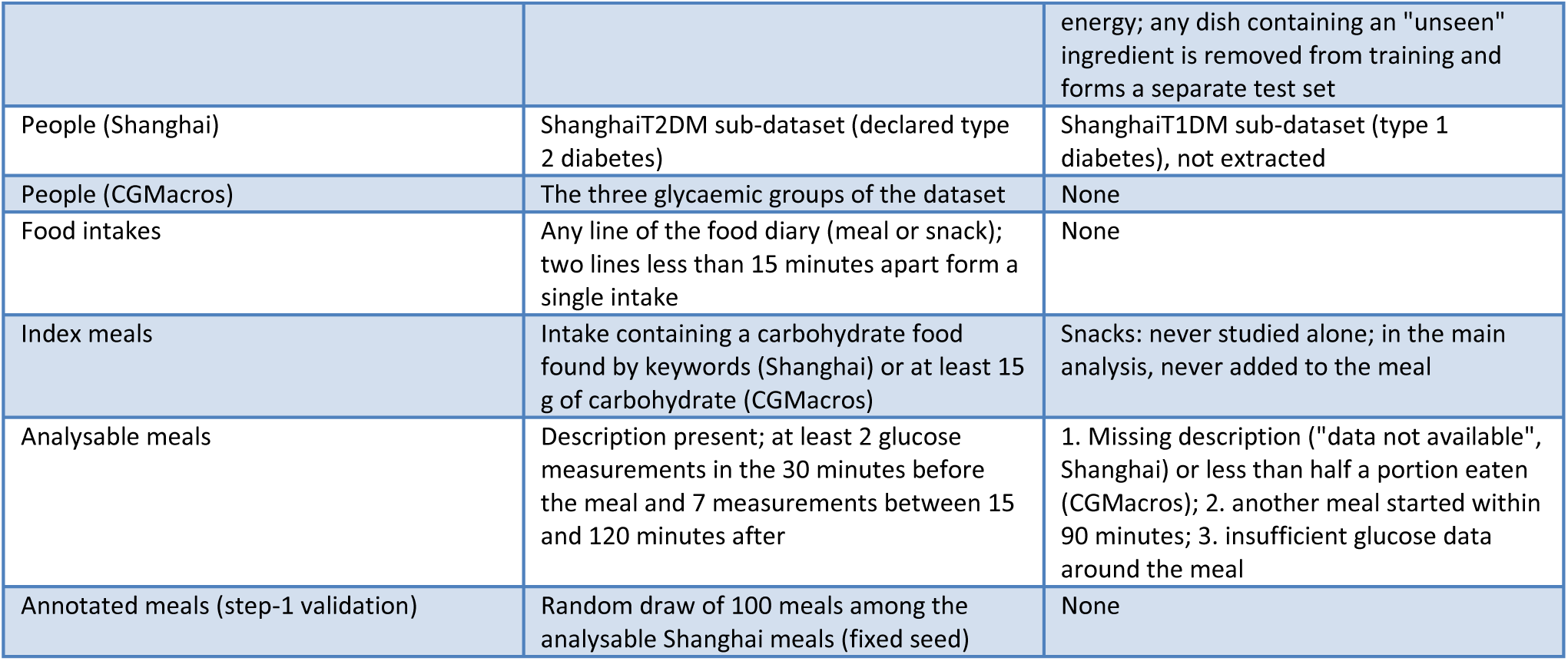
Inclusion and exclusion criteria, by level and in order of application (no counts: the numbers retained and excluded are given in the Results).

### 2.3 Glycaemic definitions

The sensors do not measure glucose at the same times: one value every 15 minutes for the Libre H in Shanghai, every 5 minutes for the Dexcom in CGMacros. To compare the two datasets, each curve was placed on the same time grid, one value every 15 minutes from meal start; in CGMacros, the value retained at each grid time was the mean of the Dexcom measurements taken within 7 minutes before or after that time. The primary outcome was **ΔGmax**, the maximum rise: the highest glucose between 0 and 120 minutes after meal start minus baseline glucose, that is, the last value before the meal. A brief peak occurring between two grid times can be missed or blunted by averaging, which may underestimate the rise; a sensitivity analysis therefore recomputed ΔGmax, in CGMacros, on the native Dexcom measurements (one every 5 minutes, without averaging).

Secondary outcomes were the 2-hour incremental area under the curve (iAUC) [28], which summarises both the height and the duration of the rise (area between the curve and the horizontal line of baseline glucose, counting only the times when glucose is above that line; trapezoidal rule), and a peak above 180 mg/dL, the upper limit of the sensors’ target range [29]. This peak depends mainly on pre-meal glucose: a person who starts a meal just below 180 mg/dL crosses it with a minimal rise. Two outcomes based on the rise itself therefore complemented it. (a) A ΔGmax rise above 50 mg/dL, whatever the starting point. Guidelines set absolute postprandial glucose targets (below 140 mg/dL two hours after the meal according to the International Diabetes Federation [30], below 180 mg/dL at peak according to the ADA [4]) but no threshold for a rise; the value of 50 mg/dL was chosen before the analyses and checked at 30 and 70 mg/dL. (b) The peak > 180 mg/dL restricted to meals starting below 140 mg/dL. Hypoglycaemia was not an outcome; sensor agreement in the low range was described (section 2.9).

### 2.4 Overview and technical details of the two models

The chain relies on two different models that share no weights (Table 3). The LLM is fine-tuned once, for a single task: converting the text of a meal into nutrients; it never sees glucose. Its outputs become, in Shanghai, input variables of a second, gradient-boosting model, trained from scratch on glucose data to predict the rise; in CGMacros, this second model receives the nutrients entered in the dataset. This two-model design was chosen from the outset; its reasons are discussed in section 4.4.

**Table 3.**
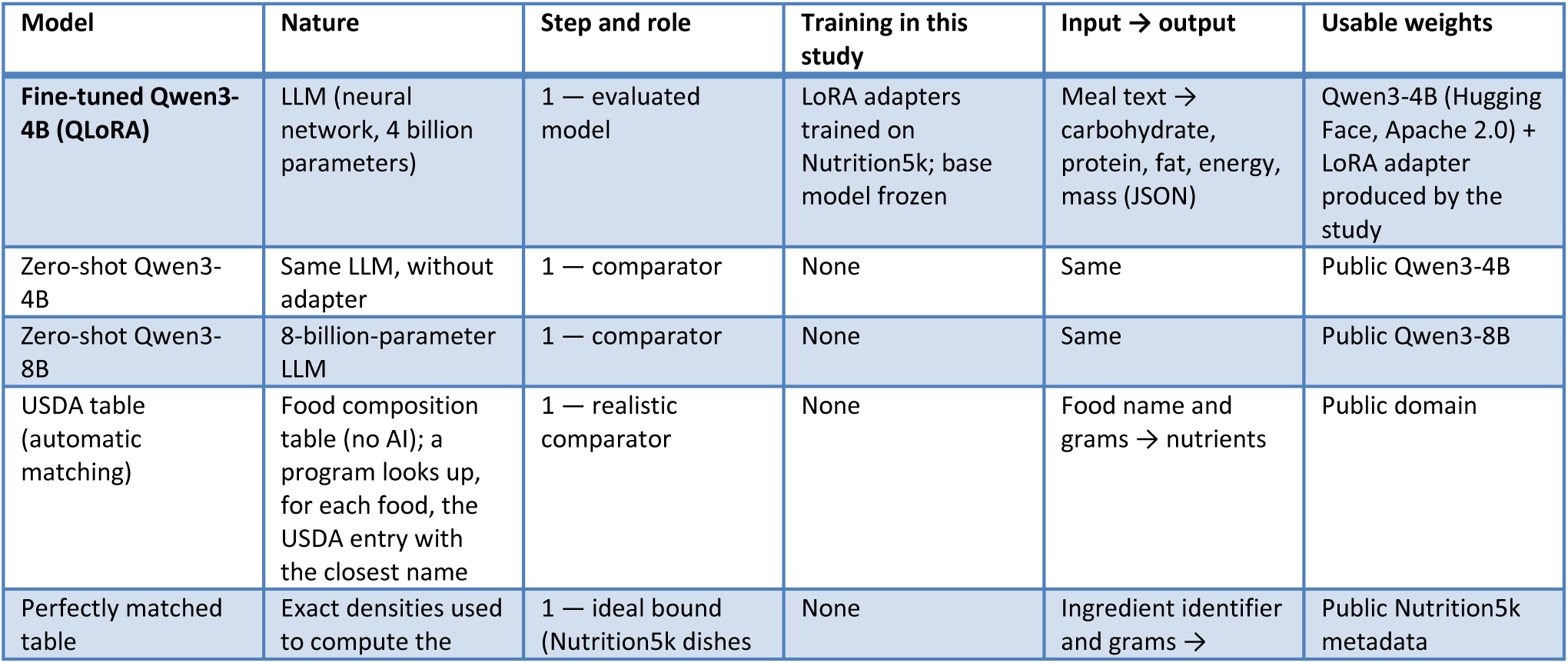
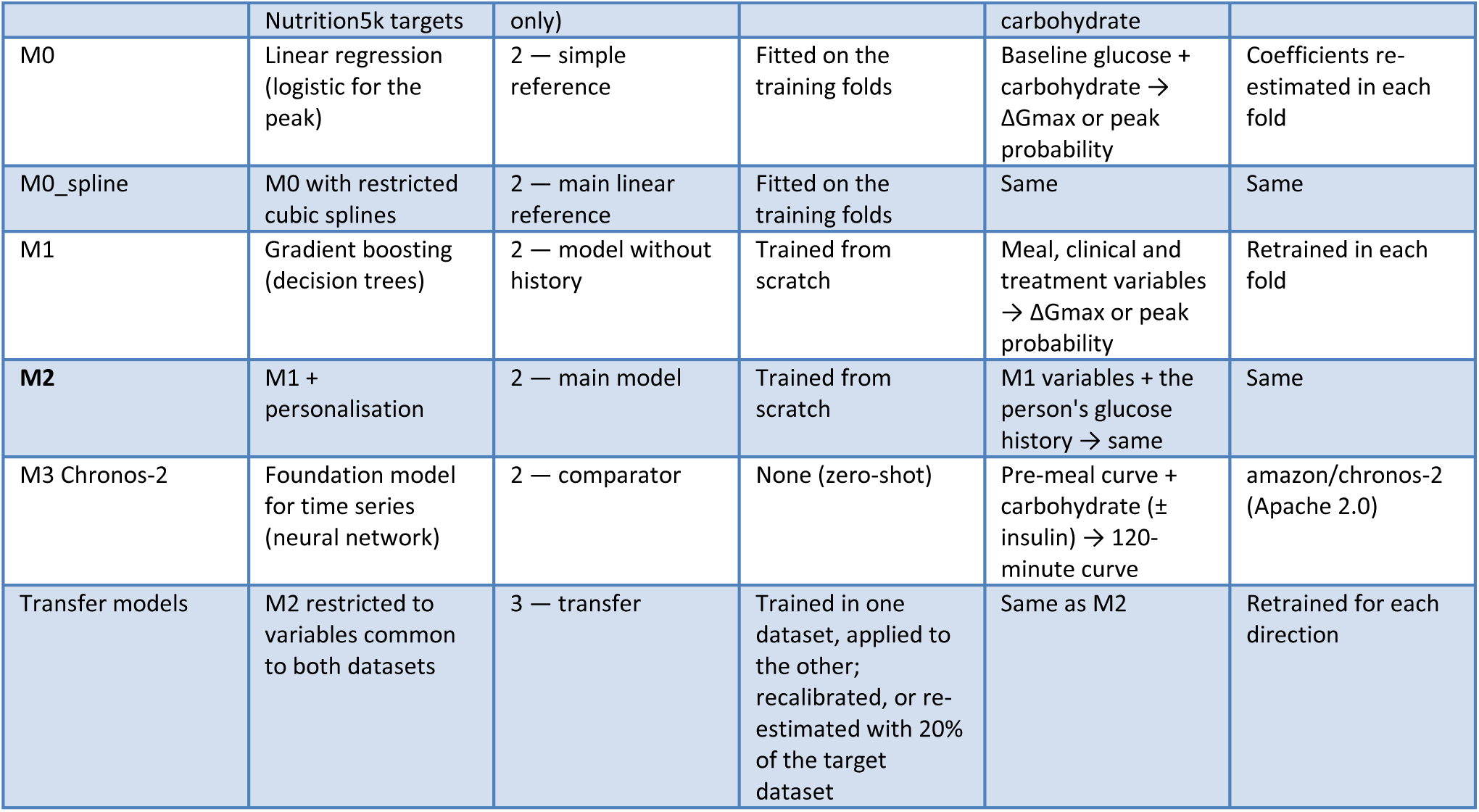
Evaluated models: nature, role, training and usable weights.

| Model | Nature | Step and role | Training in this study | Input → output | Usable weights |
| --- | --- | --- | --- | --- | --- |
| **Fine-tuned Qwen3-4B (QLoRA)** | LLM (neural network, 4 billion parameters) | 1 — evaluated model | LoRA adapters trained on Nutrition5k; base model frozen | Meal text → carbohydrate, protein, fat, energy, mass (JSON) | Qwen3-4B (Hugging Face, Apache 2.0) + LoRA adapter produced by the study |
| Zero-shot Qwen3-4B | Same LLM, without adapter | 1 — comparator | None | Same | Public Qwen3-4B |
| Zero-shot Qwen3-8B | 8-billion-parameter LLM | 1 — comparator | None | Same | Public Qwen3-8B |
| USDA table (automatic matching) | Food composition table (no AI); a program looks up, for each food, the USDA entry with the closest name | 1 — realistic comparator | None | Food name and grams → nutrients | Public domain |
| Perfectly matched table | Exact densities used to compute the | 1 — ideal bound (Nutrition5k dishes | None | Ingredient identifier and grams → | Public Nutrition5k metadata |
|  | Nutrition5k targets | only) |  | carbohydrate |  |
| M0 | Linear regression (logistic for the peak) | 2 — simple reference | Fitted on the training folds | Baseline glucose + carbohydrate → $\Delta G_{\max}$ or peak probability | Coefficients re-estimated in each fold |
| M0_spline | M0 with restricted cubic splines | 2 — main linear reference | Fitted on the training folds | Same | Same |
| M1 | Gradient boosting (decision trees) | 2 — model without history | Trained from scratch | Meal, clinical and treatment variables → $\Delta G_{\max}$ or peak probability | Retrained in each fold |
| M2 | M1 + personalisation | 2 — main model | Trained from scratch | M1 variables + the person's glucose history → same | Same |
| M3 Chronos-2 | Foundation model for time series (neural network) | 2 — comparator | None (zero-shot) | Pre-meal curve + carbohydrate (± insulin) → 120-minute curve | amazon/chronos-2 (Apache 2.0) |
| Transfer models | M2 restricted to variables common to both datasets | 3 — transfer | Trained in one dataset, applied to the other; recalibrated, or re-estimated with 20% of the target dataset | Same as M2 | Retrained for each direction |

#### The fine-tuned LLM

An LLM is a neural network trained on large amounts of text to predict the next word; Qwen3-4B has 4 billion parameters and its licence (Apache 2.0) allows local use. Fine-tuning it means continuing its training on examples of the target task, here dish descriptions paired with their nutrients. The QLoRA method makes this possible on modest hardware: the weights of the base model are stored in 4 bits instead of 16, which divides the memory needed by about four, and stay frozen; only small matrices added to each layer (the LoRA adapters) are learned (details in section 2.5). The result is a small file, the adapter, that plugs into the public base model. LLM fine-tuning and inference were run in a Kaggle session with two Tesla T4 graphics processing units (GPUs); the 4-bit model fits in the memory of a consumer graphics card, but running it on a personal computer or a phone was not evaluated.

#### Gradient boosting

Gradient boosting [31, 32] builds a sequence of shallow decision trees; each tree is a series of simple questions on the variables (“is pre-meal glucose above a given value?”, “does the meal contain more than a given amount of carbohydrate?”) that ends in a predicted value, and each new tree corrects the remaining error of the previous ones. The method handles missing values and non-linear relationships. The implementation was scikit-learn’s HistGradientBoosting [33], with settings fixed before the analyses: a learning rate of 0.05, meaning that each tree contributes only 5% of its correction (the scikit-learn default is 0.1; a lower rate, offset by more trees, limits overfitting [31]); an L2 regularisation of 1, a penalty that moderates extreme leaf values. Three settings were chosen automatically: the number of leaves per tree (15 or 31), which sets the complexity of each tree; the minimum number of meals per leaf (20 or 50), which prevents rules based on too few meals; and the number of trees (150 or 300). This choice used nested person-level cross-validation: within the training data only, people were split again into subgroups to compare the combinations, so that test people were never used to choose the settings. For the peak, a separate classifier of the same family provided a probability. Gradient boosting, the linear references and Chronos-2 were run on an ordinary computer, on its central processing unit (CPU), without a graphics card.

### 2.5 Step 1: nutrient estimation by LLM

#### Splits

Ten per cent of distinct ingredients, drawn at random outside the 50 most frequent (seed 42), were declared “unseen”: any dish containing at least one formed the “unseen ingredients” test set and was removed from training. The other dishes were split by dish: 80% for training, 10% for validation (monitoring of training) and 10% for testing. Training texts were augmented (shuffled ingredient order, 20% of dishes described in portions rather than grams, case and plural variations). A “portions” variant of the test set evaluated descriptions without grams. Test dishes whose set of ingredients appears in no training dish (“new combinations”) were also evaluated separately. Three sets were therefore held out from training, from closest to furthest: test dishes, new combinations and dishes with unseen ingredients; to these were added portion descriptions and, as external validation, the 100 real Shanghai meals.

#### Fine-tuning (QLoRA)

The base model was Qwen3-4B [34], in its public version on Hugging Face at training time; the adapter produced is identified by its SHA-256 hash (supplementary material). It was fine-tuned with QLoRA [35]: the base model weights, quantised to 4 bits (NF4 format, double quantisation), stayed frozen, and only LoRA adapters [36] added to all linear layers were trained, in 16-bit precision. Each adapter consists of two small matrices whose product corrects the layer output; their rank (16) sets their size, and the factor α = 32 the amplitude of the correction. The loss applied only to the expected answer, not to the question. Hyperparameters were fixed before training: learning rate 0.0002, decreased progressively along a cosine curve; at most 2 epochs (complete passes over the training dishes); early stopping if the validation loss stopped decreasing (patience of 3 evaluations, one evaluation every 20 steps); planned effective batch of 32, that is, 32 examples seen before each weight update (2 examples × 16 accumulation steps on one GPU). As the two GPUs of the session were used in parallel, the effective batch was doubled (protocol deviation). With this number of steps, early stopping could only trigger at the very end of training.

#### Comparators: two composition tables, two uses

The comparators were Qwen3-4B and Qwen3-8B used as they are, without examples or training (“zero-shot”), and two food composition tables, not to be confused.

- The **automatically matched USDA table** is the realistic comparator: it reproduces what nutrition software without AI would do. For each food in the text, a program looks up the USDA entry with the closest name (string similarity, without any hand-chosen correspondence), then multiplies its carbohydrate content by the stated grams; if no name is close enough, it imputes the median content of the table. It can therefore pick the wrong food, or the wrong state of a food (raw or cooked, fresh or dried), and this is the error the LLM must avoid.
- The **perfectly matched table** is not a realistic comparator but an ideal bound, computable only on Nutrition5k dishes. For each ingredient, it uses the carbohydrate content published by Nutrition5k for that exact ingredient, the very one used to compute the dish’s target carbohydrate, and multiplies it by the grams written in the text. It thus redoes the computation that produced the target: its error is almost zero by construction and comes only from the rounding of the grams written in the text. It answers the question: “what would a table do if every food were recognised without error?”. It does not exist for Shanghai meals, whose foods are not in Nutrition5k.

At inference, all LLMs were loaded in 4 bits, received the same prompt (written instruction), frozen and versioned (full text in the supplement), without reasoning mode, and decoded greedily: at each step the most probable word is kept, without random sampling, so that the same text always gives the same answer. The output had to be a JSON object, a standard “key: value” text format readable by a program (for example “glucides_g: 30”), containing carbohydrate, protein, fat, energy and mass, validated against a schema; an invalid output triggered a second attempt, then was counted as missing. As the LLM returns only nutrients, not a list of foods, invented foods (hallucinations) could not be counted.

#### Measures

The mean absolute error (MAE: mean, over all dishes, of the gap in grams between estimated and reference carbohydrate, regardless of sign) and the relative MAE (the same gap divided by the reference value), per nutrient; the proportion of dishes estimated within ±10 g of carbohydrate; Lin’s concordance coefficient [37], which equals 1 if the estimates coincide exactly with the reference; the bias (mean signed gap) and the Bland-Altman limits of agreement [38], the interval containing 95% of the gaps; the rate of valid JSON; and the proportion of answers whose energy differed by more than 25% from 4 × carbohydrate + 4 × protein + 9 × fat (Atwater factors, in kcal per gram). This last check verifies the internal consistency of each answer: an energy incompatible with the stated nutrients signals an invented rather than computed answer. Error was also described by the number of ingredients in the dish (1–2, 3–6, 7 or more). On real meals, paired MAE differences were added (the error difference between two estimators on the same meals, which neutralises the difficulty specific to each meal), with CIs computed by resampling meals (meal-clustered CI), as well as the median relative error and a regression-based Bland-Altman analysis, in which the bias and the width of the limits can vary with meal size [39]. **Pass criterion** set in advance: the fine-tuned model had to beat the best zero-shot model on carbohydrate MAE, the CI of the difference excluding 0; otherwise the best untrained reference would have been used in step 2.

#### Validation on real meals: the reference

To judge an estimate, a reference value as close as possible to the true amount of carbohydrate is needed. For the 100 randomly drawn Shanghai meals, this reference was computed from the quantities weighed and recorded by the patients, with the USDA table, by a Python program written for the study; it serves as the common yardstick for all compared estimators (fine-tuned LLM, zero-shot LLMs, automatically matched table). This program performs no automatic matching: it applies an explicitly written correspondence table, linking each food in the text to one or more named USDA entries, chosen with the help of the original Chinese text (米饭, cooked rice, versus 米, raw rice; 馒头, plain steamed bun, versus 包子, stuffed bun), and it breaks composite dishes down into recipes (mass proportions). A reference is never 100% exact: some Chinese dishes have no exact equivalent in the American table and the recipe of a composite dish is estimated. Each reference was therefore given a certainty level: 1 = approximate (food or recipe approximated), 2 = probable, 3 = certain (simple food present in the table). The investigator (physician, first author, not blinded to the hypothesis) checked each line without access to the outputs of the evaluated LLMs. The 100 meals, all certainty levels combined, form the main analysis; error was also computed on certainty-2 or -3 meals only, to check that the result does not depend on the most uncertain references.

In a sensitivity analysis, the reference was recomputed by a second, deterministic program that reads the original Chinese text rather than the English translation (the quantities of the two versions sometimes differ), with its own correspondence table, written from the Chinese labels, and lower and upper bounds for each food (cooked or raw, dough share, added sugar). This recomputation is not a prediction model but a check: if the errors measured against the two references are close, the conclusion depends neither on the translation nor on the matching choices. It uses the same USDA table and therefore does not constitute an independent annotation.

### 2.6 Step 2: prediction of the glucose rise

#### Per-meal variables

Baseline glucose; 30-minute slope, that is, the rate of change of glucose (mg/dL per minute) in the half hour before the meal, which indicates whether it was already rising or falling; variability over the previous 3 hours; time of the meal, coded by its sine and cosine, two coordinates on a clock circle that place late evening and early night side by side, as on a dial; rank of the meal in the day. Nutrients: carbohydrate, protein, fat, energy and carbohydrate share of energy (estimated by the selected LLM in Shanghai, entered in CGMacros); protein and fat slow gastric emptying and blunt the rise after a carbohydrate meal, including in T2D [40]. Clinical variables: age, sex, BMI, HbA1c, diabetes duration (Shanghai) or fasting glucose (CGMacros). In Shanghai: treatment classes, prandial insulin dose and intake with the meal (-30 to 0 min in the main analysis) of an alpha-glucosidase inhibitor (AGI, such as acarbose, which slows carbohydrate digestion) or of a secretagogue (sulfonylurea or glinide, which stimulates insulin secretion). When energy was incompatible with carbohydrate (4 × carbohydrate > energy, i.e. a carbohydrate share above 1), it was recomputed as 4 × carbohydrate + 4 × protein + 9 × fat.

The available variables differ between datasets (diabetes duration exists only in Shanghai, fasting glucose only in CGMacros). As each model is developed and validated within a single dataset, with that dataset’s variables, this difference introduces no bias in the evaluation of each dataset; it does, however, limit direct comparisons. This is why the between-dataset comparison (harmonised analysis) and the transfer (step 3) use only the variables common to both datasets.

The prandial insulin dose is chosen according to the planned meal: it partly reflects its carbohydrate content (it is an indicator of it) and it reduces the rise (it lies on the path between meal and glucose, like a mediator). Including this dose may therefore capture part of the effect of carbohydrate; a sensitivity analysis retrained M2 without it.

#### Missing data

Missing values are expected in routine-care data; their frequency by variable is reported in the Results, as required by TRIPOD+AI [1]. Gradient boosting handles them natively: at each question of a tree, meals without a value are sent to the branch that most reduces the error. For linear models, a missing value was replaced by the median of the variable, and variables were standardised, both operations being learned on the training fold only (scikit-learn pipeline). Meals excluded for missing description were compared with included meals (pre-meal glucose, ΔGmax, time, prandial insulin).

#### Personalisation

For each meal, the model receives the person’s usual rise, computed only from their **previous** meals in the dataset (those whose rise had already been measured by the sensor): the mean of the ΔGmax already observed in that person, shrunk towards the population mean of ΔGmax according to (sum of previous rises + k × population mean) / (number of previous meals + k), together with the number of meals already observed. This formula is the classical shrinkage estimator, or “credibility mean”, of empirical Bayes statistics [41, 42]: with few meals, the value stays close to the population mean; as meals accumulate, it moves towards the person’s own mean. The parameter k acts as a number of fictitious meals equal to the population mean. It was set to 3 before the analyses (“fixed-weight shrunk mean”): the personal mean weighs as much as the population mean from the third observed meal onwards, a moderate shrinkage. The population mean is learned on the training folds only, and no information from the current or future meals is used. Credibility theory also provides an estimated value of k, the ratio of the within-person to the between-person variance of rises, computed here by the method of moments (from the variances observed in the training folds) [42]; sensitivity to the choice of k was examined (k ∈ {1; 3; 10; estimated k}), and the “personal mean alone” (the usual rise used directly as the prediction) served as a comparator.

#### Compared configurations

Variable families were added one at a time (Table 4); level 1 already contains time and meal rank, so the carbohydrate gain measured at level 2 is the gain once these variables are known. In Shanghai, to find out whether a more accurate carbohydrate count improves the prediction of the rise, M2 was retrained with each nutrient source: fine-tuned LLM, zero-shot LLM (4B, 8B), USDA table, or a plain sum of grams (total grams, grams of carbohydrate foods and number of foods, without any conversion into nutrients). M3, Chronos-2 [43, 44], a model pre-trained by Amazon on a very large number of time series, was used as it is: it received the glucose curve of the day preceding the meal and carbohydrate (and insulin in Shanghai) as covariates, and extended the curve over 120 minutes, from which ΔGmax and the peak were read. Two linear references used baseline glucose and carbohydrate: M0, in which they enter as they are (the predicted rise varies in a straight line with each), and M0_spline, in which they enter through restricted cubic splines with 5 knots [45]. A spline divides the scale of a variable into intervals and fits a piece of curve on each, joined smoothly to its neighbours: the model remains a linear regression, but the relationship between starting glucose (or carbohydrate) and the rise can be curved. M0_spline is the main linear reference. To understand a possible weakness of M0, diagnostics were planned: M0 with glucose alone, a linear model on the level-1 variables, gradient boosting with glucose alone or with glucose and carbohydrate, the standardised coefficients (change in the predicted rise for a one-standard-deviation change in the variable), the correlation obtained in each of the five folds, the number of imputed values, and the relationship between starting glucose and the rise, between people (people’s means) and within people (deviations from each person’s mean). Finally, a “leave-one-family-out” ablation removed one family at a time from M2 (pre-meal glucose; time and rank; nutrients; clinical; treatment; personalisation), with paired CIs and Holm correction.

**Table 4.** Step-2 configurations: each level adds a family of variables to the previous one.

| Level | Name | Added variables | Use |
| --- | --- | --- | --- |
| 1 | Pre-meal glucose | Baseline glucose, 30-min slope, 3-h variability, time, meal rank | Ablation; "glucose only" reference |
| 2 | + carbohydrate | Meal carbohydrate (or raw grams in Shanghai, "grams" source) | Ablation |
| 3 | + macronutrients | Protein, fat, energy, carbohydrate share | Ablation; comparison of sources |
| 4 | + clinical (and treatment in Shanghai) | Age, sex, BMI, HbA1c, diabetes duration or fasting glucose; treatment classes and intakes with the meal (Shanghai) | **M1** |
| 5 | + personalisation | Shrunk usual rise and number of previous meals | **M2 (main model)** |
| 6 | + watch (CGMacros) | Pre-meal heart rate (and post-meal, in sensitivity) | Secondary analysis |

#### Validation and measures

The people of each dataset were randomly divided into five groups of similar size, the **folds**, keeping in each fold the proportion of each glycaemic group (CGMacros) or of patients on insulin (Shanghai): this is stratification. Each fold serves once as the test set, the model being trained on the other four (the training folds). A person therefore never appears in both training and testing, and each meal receives a prediction made by a model that has never seen that person: this is the **out-of-fold prediction**, on which all analyses are based. For ΔGmax: Pearson correlation r between observed and predicted values, MAE (mg/dL), calibration slope and intercept, and, as co-primary error measures, RMSE, out-of-fold R², skill score (1 - model MAE / MAE of a null model: training-fold population mean, then previous personal mean) and within-person correlation of out-of-fold predictions (mean of per-person r among people with at least 8 meals). For the peak: AUROC, Brier score and calibration slope. Each measure is defined in Box 1. M2 performance was described by subgroup: glycaemic group (CGMacros), treatment class of the recording (Shanghai), sex (both datasets) and self-reported origin (CGMacros). The decision curve of the “peak > 180 mg/dL” alert [46] was described over the whole range of thresholds, with bootstrap CIs and the net-benefit difference M2 - M0_spline (ΔNB) over thresholds 0.2 to 0.8; as no alert threshold is established in clinical practice, the 0.5 threshold is only a reading point. Repeated 5 × 5 cross-validation (five person-level splits, seeds 42 to 46) described the variability of r, MAE, AUROC and the carbohydrate gain.

###### Box 1. Definitions of measures and statistical terms

Box 1. Definitions of measures and statistical terms.
| Term | Definition |
| --- | --- |
| MAE (mean absolute error) | Mean of the gaps, regardless of sign, between estimated and reference value (g of carbohydrate, or mg/dL of rise); 0 = no error |
| Bias | Mean of the signed gaps: negative if the model underestimates on average |
| Bland-Altman | Plot of the gaps against the mean of the two measurements; limits of agreement = bias ± 1.96 standard deviations, the interval containing 95% of the gaps. Regression-based version: the bias and the width of this interval can vary with meal size |
| Lin's concordance coefficient | Measure of agreement between two series of values, from 0 to 1; 1 = identical values |
| Paired difference | Difference in error between two methods computed on the same dishes or the same meals |
| Pearson correlation r | Degree of alignment between predicted and observed rises, from -1 to 1: 0 = no relationship, 1 = predictions perfectly aligned with reality. For the postprandial glycaemic response, published personalised models reach $r = 0.62$ [12] to $0.69$ [21] |
| Spearman correlation $\rho$ | Same principle as r, computed on ranks; less sensitive to extreme values |
| Within-person correlation | Correlation computed separately in each person, then averaged: does the model distinguish the meals of the same person? |
| Out-of-fold $R^2$ | Share of the variability of observed rises reproduced by the predictions (0 to 1) |
| RMSE | Square root of the mean squared error; penalises large errors more than the MAE |
| Skill score | $1 - \text{model MAE} / \text{MAE of a simple comparison model}$; positive if the model makes less error than this simple model |
| AUROC | Probability that a meal followed by a peak receives a higher predicted probability than a meal without a peak; 0.5 = chance, 1 = perfect discrimination |
| Brier score | Mean of the squared gaps between predicted probability and observed event (0 or 1); the lower, the more accurate the probabilities |
| Calibration slope | 1 = predictions on the right scale; $< 1$ = predictions too spread out |
|  | (too high for large rises, too low for small ones) |
| Net benefit (decision curve) | Gain of an alert: proportion of true positives minus proportion of false positives weighted by the decision threshold; compared with "always alert" and "never alert" |
| 95% CI by cluster BCa bootstrap | Confidence interval obtained by drawing at random, with replacement, whole people (or meals), then correcting for bias and skewness of the distribution (BCa method) |
| Holm correction | Adjustment of p values when several comparisons are made, to limit false positives |
| Person-level cross-validation, folds, out-of-fold prediction | See section 2.6: each prediction comes from a model that has never seen the person concerned |

#### Between-dataset comparison and harmonised analysis

The difference in carbohydrate gains (CGMacros - ShanghaiT2DM), for r, MAE and R², was estimated by independent bootstrap within each dataset (people drawn with replacement, 2,000 resamples; one seed per dataset, 42 and 43, so that the two resamplings are independent), with a two-sample BCa CI (acceleration by jackknife over the people of both datasets) and a two-sided bootstrap p value. The harmonised analysis used the level-1 variables, present in both datasets, and meals with at least 15 g of carbohydrate, in the CGMacros people with T2D on the one hand, and in Shanghai on the other hand in two subsets: meals eaten without prandial insulin, AGI or secretagogue, then people without insulin or secretagogue. In Shanghai, sensitivity analyses excluded the recordings with an insulin pump (continuous subcutaneous infusion) and those whose “intravenous insulin” column was filled; this column records occasional infusions of fluid with added rapid-acting insulin, given in addition to subcutaneous or pump insulin. On the 100 annotated meals, the relationship between ΔGmax and reference carbohydrate was compared with that obtained with LLM carbohydrate (Spearman correlation and slope), to describe the attenuation due to measurement error.

#### Sample size

TRIPOD+AI requires the sample size to be justified [1]. It was set here by the available public datasets; its adequacy was checked against the minimum sample size required to develop a model with a continuous outcome, computed according to Riley et al. [47, 48], with the formulas of the pmsampsize software [49] reimplemented in Python: expected shrinkage of at least 0.9, a difference of at most 0.05 between apparent and adjusted R², precise estimation of the residual standard deviation and of the mean (multiplicative margin of 1.1), using the observed out-of-fold R² of M2 and the number of candidate variables. The calculation ignores the clustering of meals within people.

### 2.7 Step 3: transfer between datasets

As not all variables are shared by the two datasets, transfer models used only the variables present in both. Models trained in CGMacros (T2D and prediabetes, then the whole dataset) were applied to ShanghaiT2DM, and conversely (to the CGMacros people with T2D, then the whole dataset); one variant used the CGMacros Libre Pro, of the same sensor family as the one in Shanghai (six scenarios). An affine recalibration corrected the scale of predictions with a straight line (corrected prediction = a + b × initial prediction), learned on 20% of the people of the target dataset, evaluation being done on the remaining 80%. Such an increasing correction does not change the order of meals, hence neither r nor the AUROC: only calibration and MAE can change. An update by re-estimation retrained the model on the source dataset plus the 20% of target people [50]. The internal reference (same set of variables, 5 person-level folds in the target dataset) and full M2 were evaluated on the same 80% of people.

### 2.8 Complementary (exploratory) analyses

(A) Relationship between carbohydrate and ΔGmax: coefficient of variation of carbohydrate per meal (standard deviation divided by the mean, which measures their spread), overall and within-person Spearman correlation (mean of per-person correlations among people with at least 8 meals), in each dataset, in the CGMacros people with T2D and, in Shanghai, for meals eaten without prandial insulin or AGI, with the carbohydrate gain in these subsets. (B) Grams per food category extracted from the text (16 categories) added to M2 in Shanghai. (C) Variable importance of M2 by permutation [51]: in the test meals, the values of a family of variables are shuffled at random between meals, which breaks their link with the rise, and the increase in error (MAE) is measured; the larger the increase, the more the model relied on that family. All variables of a family were shuffled together, so as not to split importance between correlated variables. Spearman correlations between carbohydrate and time, rank, starting glucose and prandial insulin dose were added. (D) Decision curve (section 2.6). (E) Mean glucose curves by carbohydrate tertile (three groups of equal size, from the third of meals with the least carbohydrate to the third with the most), with tertile bounds and person-clustered CIs.

### 2.9 Sensitivity analyses and sensor agreement

The exclusion window for close meals was changed from 90 to 60 and then 120 minutes, and the Libre Pro replaced the Dexcom as the CGMacros sensor. The analysis with information up to +30 min and +2 h (section 2.1) was compared with the main analysis. In the same CGMacros people, episodes below 70 mg/dL for at least 15 minutes were counted on each of the two sensors; in Shanghai, low sensor readings were compared with capillary glucose values taken within ±15 minutes.

### 2.10 Statistics

95% CIs by cluster BCa bootstrap [52]: units are drawn at random with replacement, 2,000 times, and the computation is repeated at each draw; the unit drawn is the person for steps 2 and 3 (all the meals of a person stay together) and, for step 1, the dish (Nutrition5k) or the annotated meal (Shanghai); seed 42. Paired comparisons on the same meals, with a two-sided bootstrap p value and Holm correction [53] within each family of comparisons (ablations; nutrient sources; complementary analyses; leave-one-family-out). Calibration: slope and intercept (regression) or slope and calibration-in-the-large (logistic) [54]. Python 3.13.5, scikit-learn 1.8.0, pandas 2.3.3. Every number in this manuscript is generated automatically from the pipeline outputs: no value is typed by hand.

## 3. Results

### 3.1 Populations, dishes and meals

**Figure 1.**
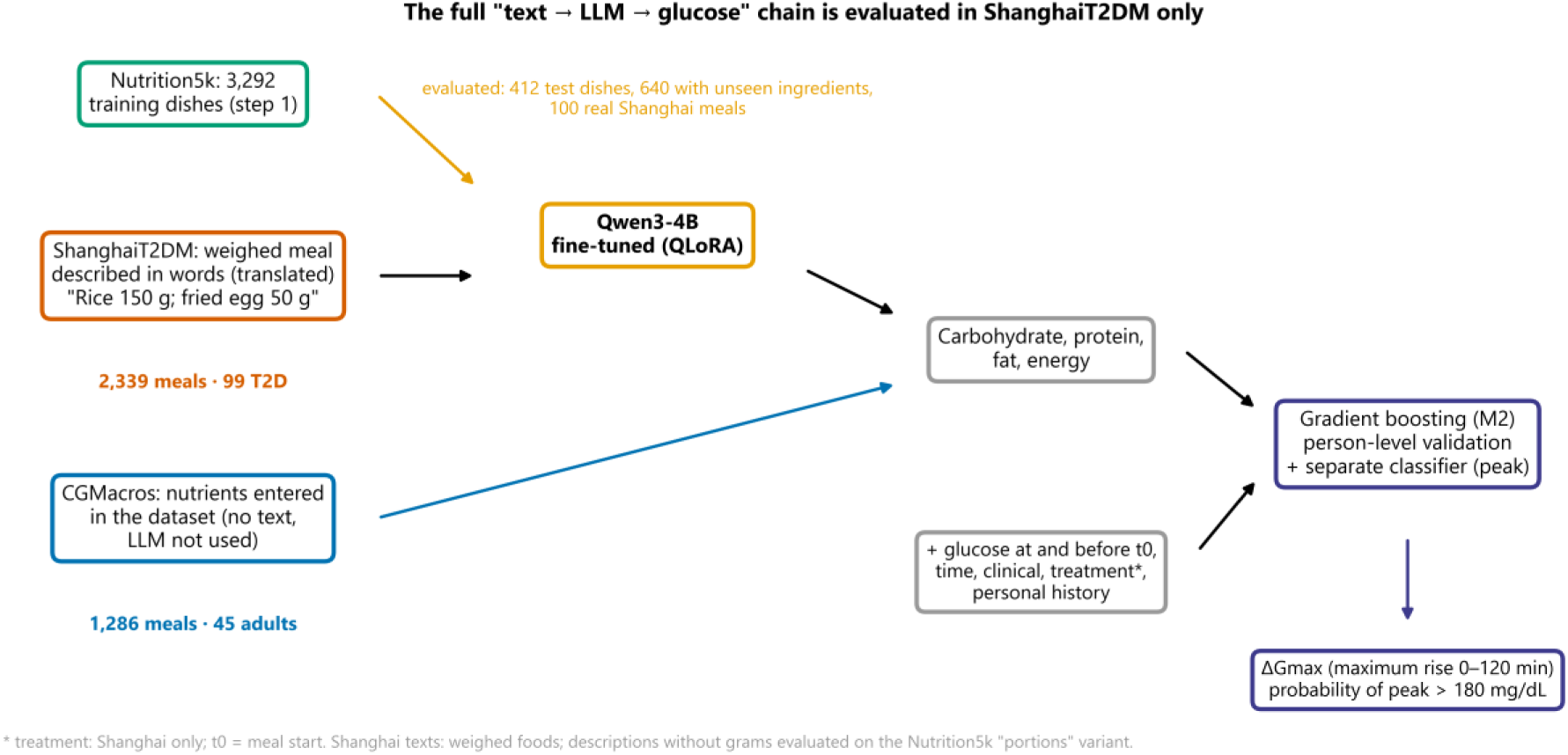
Study pipeline: two input branches (ShanghaiT2DM: meal text read by the LLM; CGMacros: nutrients entered in the dataset) and numbers at each step.

Nutrition5k contained 4,768 dishes, of which 13 were excluded for inconsistency (11 whose total mass differed by more than 20% from the sum of the ingredient masses, 2 with zero energy), leaving 4,755 dishes (210 distinct ingredients, 2,305 ingredient combinations). The draw designated 21 unseen ingredients, present in 640 dishes; the 4,115 other dishes were split into 3,292 for training, 411 for validation and 412 for testing.

The Shanghai archive contained 112 people: the type 1 diabetes sub-dataset (12 people, 16 recordings) was not extracted. The 100 people with T2D had 109 recordings; 35 of them (32%) included pump insulin. The “intravenous insulin” column was filled in 6 recordings: 5 described 1 to 5 infusions of fluid with added rapid-acting insulin, all in patients also receiving subcutaneous or pump insulin, and 1 contained the heading of another column. The 3,693 food intakes included 621 snacks (34 eaten within 2 hours after an analysable meal, 587 not), leaving 3,072 meals; 733 were excluded, including 663 without description (22% of meals), 37 followed by another meal within 90 minutes and 33 with insufficient glucose data, leaving 2,339 analysable meals in 99 people (1 person without any analysable meal; 20 [8, 33] meals per person). For the 636 meals excluded for missing description whose glucose outcomes were computable, median pre-meal glucose was 124 mg/dL, ΔGmax 31 mg/dL and median time 11.6 h, versus 128 mg/dL, 45 mg/dL and 11.5 h for included meals; prandial insulin accompanied 39% and 24% of these meals.

CGMacros contained 45 people (15 normoglycaemic, 16 prediabetic, 14 T2D; no type 1 diabetes; 34 self-reporting Hispanic or Latino origin). The 1,608 intakes included 185 snacks (28 eaten within 2 hours after an analysable meal), leaving 1,423 meals; 137 were excluded (8 for less than half a portion eaten, 84 for another meal within 90 minutes, 45 for insufficient glucose data), leaving 1,286 analysable meals in 45 people (Figure 2). Energy was recomputed for 31 CGMacros meals (carbohydrate share up to 3.30 before correction) and 37 Shanghai meals (up to 1.13). Moving to strict t0 changed the predictors of 28 CGMacros meals and 34 Shanghai meals (snacks) and the treatment of 101 Shanghai meals. The median rise was 45 mg/dL in Shanghai and 59 mg/dL in the CGMacros people with T2D; a peak above 180 mg/dL followed 48% and 66% of meals, respectively.

**Figure 2.**
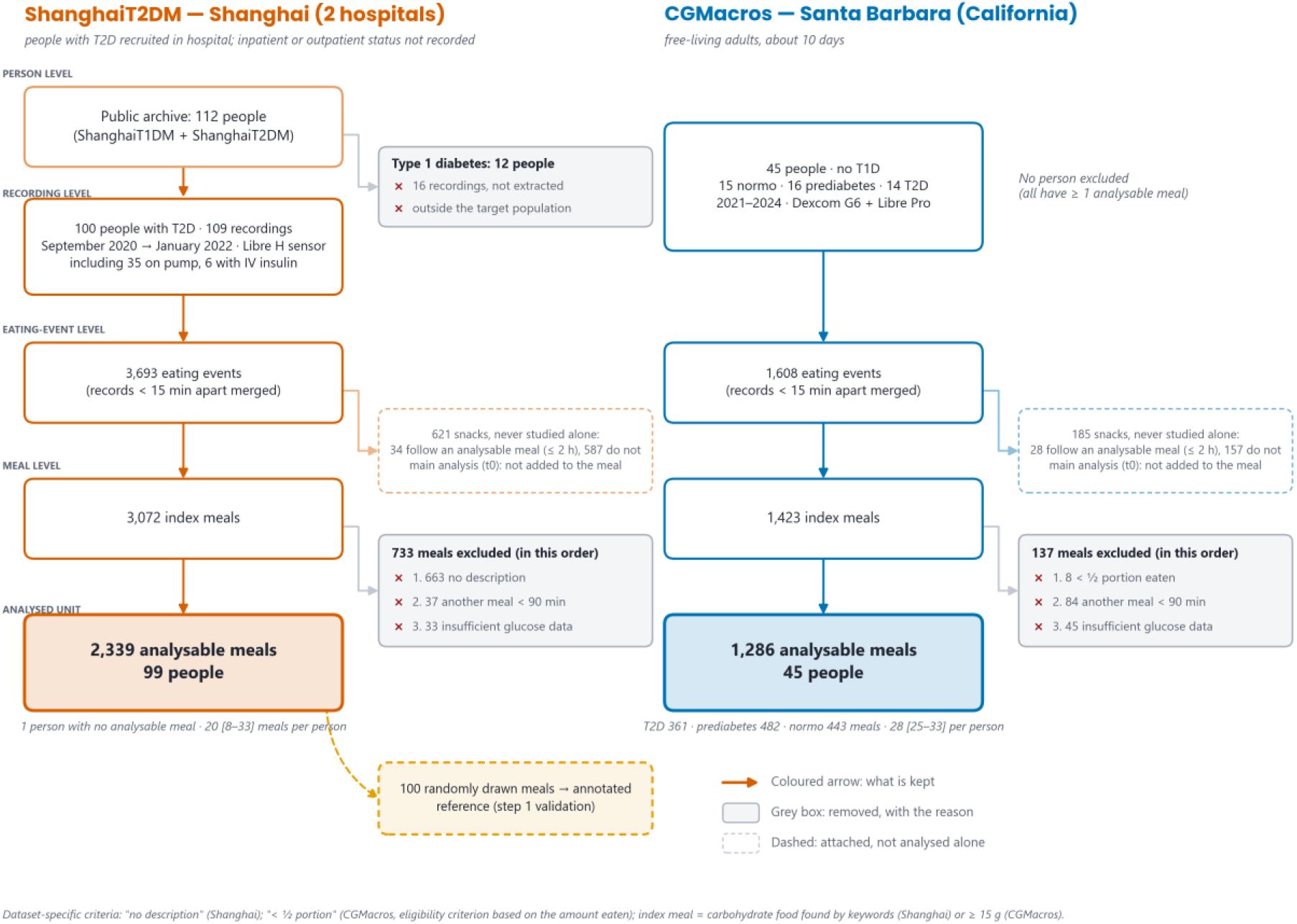
Flow diagram by level (person, recording, intake, meal): at each level, the elements excluded, in order of application, with their reason; fate of snacks; recordings with pump or intravenous insulin; meals per person.

Among analysable meals, 90% of Shanghai meals and 97% of CGMacros meals had all their variables recorded. The most frequently missing variable was HbA1c in Shanghai (9.5% of meals) and pre-meal heart rate in CGMacros (3.4%); glucose variables, nutrients, age, sex and BMI were missing for no meal.

**Table 5.** Characteristics of participants and meals (median [IQR] or %).

|  | Shanghai (T2D) | CGMacros, all | CGMacros, T2D |
| --- | --- | --- | --- |
| People | 99 | 45 | 14 |
| Analysable meals | 2,339 | 1,286 | 361 |
| Meals per person | 20 [8, 33] | 28 [25, 33] | 25 [24, 28] |
| Age (years) | 63 [51, 68] | 51 [40, 58] | 52.5 [51.0, 59.0] |
| Women (%) | 44 | 64 | 57 |
| BMI (kg/m<sup>2</sup>) | 23.8 [22.2, 25.5] | 30.0 [26.9, 35.9] | 30.6 [28.2, 34.3] |
| HbA1c (%) | 8.5 [7.1, 11.0] | 5.9 [5.5, 6.9] | 7.1 [6.9, 7.4] |
| Pre-meal glucose (mg/dL) | 128 [101, 155] | 119 [105, 141] | 141 [116, 169] |
| Carbohydrate per meal (g) | 30 [20, 44] | 66 [29, 76] | 66 [34, 73] |
| Diabetes duration (years) | 7 [1, 14] | Not recorded | Not recorded |
| Insulin / metformin / AGI / secretagogue (%) | 49 / 42 / 25 / 17 | Not recorded | Not recorded |

Shanghai carbohydrate: estimated by the fine-tuned LLM; CGMacros: entered in the dataset. CGMacros records neither diabetes duration nor per-meal treatments.

### 3.1 Step 1: nutrient estimation

The step-1 results follow the order of the pipeline: LLM training, internal control on Nutrition5k, then external validation on the 100 real Shanghai meals. Table 6 summarises them; its seven rows correspond, from top to bottom, to the evaluation sets from closest to furthest from training: Nutrition5k test dishes; test dishes with a new ingredient combination; dishes with unseen ingredients; portion descriptions; the 100 real Shanghai meals; these same meals restricted to certainty-2 or -3 references; and the proportion of test dishes estimated within ±10 g.

**Table 6.** Carbohydrate estimation: MAE in grams [95% CI], from the sets closest to training to the furthest.

| Set | Fine-tuned Qwen3-4B (QLoRA) | USDA table (automatic matching) | Zero-shot Qwen3-4B | Zero-shot Qwen3-8B |
| --- | --- | --- | --- | --- |
| Nutrition5k test dishes (n = 412) | **2.2 [2.0, 2.5]** | 8.3 [6.7, 10.8] | 14.5 [12.4, 17.3] | 17.8 [15.4, 20.7] |
| Test, new combinations (n = 166) | **3.1 [2.7, 3.6]** | 9.4 [7.0, 13.2] | 21.9 [17.8, 27.4] | 26.0 [21.3, 31.8] |
| Unseen ingredients (n = 640) | **4.8 [3.8, 8.4]** | 8.5 [7.4, 10.2] | 30.6 [27.3, 34.6] | 35.2 [32.2, 39.1] |
| Portion descriptions (n = 412) | **4.0 [3.6, 4.4]** | Not computable (no grams) | 11.4 [9.9, 13.5] | 15.9 [13.7, 18.6] |
| Real Shanghai meals (n = 100) | **12.2 [9.9, 15.2]** | 22.6 [18.0, 28.7] | 34.3 [29.3, 40.2] | 33.1 [26.6, 42.9] |
| Real meals, certainty 2-3 (n = 77) | **12.0 [9.5, 15.9]** | 23.2 [17.9, 30.6] | 32.5 [27.0, 39.2] | 28.2 [22.1, 36.7] |
| Test dishes within $\pm 10$ g | 97% | 85% | 67% | 60% |

#### Training

Training comprised 104 steps and 33.0 million trained parameters; the validation loss went from 0.678 at the first evaluation to 0.546 at the best checkpoint, reached at the last step. The effective batch was 64 instead of 32. The fine-tuned model produced valid JSON in 100.0% of cases, with no missing value in any set.

#### Internal control: Nutrition5k dishes

On test dishes, for which the grams of each ingredient are given, the carbohydrate MAE of the fine-tuned model was 2.2 [2.0, 2.5] g, with 97% of dishes within ±10 g and a Lin concordance of 0.97. The pass criterion was met: MAE difference with the best zero-shot model of -12.3 [-14.9, -10.3] g (p < 0.001). The MAE difference with the automatically matched USDA table was -6.1 [-8.5, -4.5] g on the test set and -3.8 [-5.1, -1.8] g on unseen ingredients. Of the 412 test dishes, 166 had an ingredient combination absent from training: the MAE of the fine-tuned model was 3.1 [2.7, 3.6] g on these, versus 1.6 [1.3, 2.0] g for seen combinations. On portion descriptions, it was 4.0 [3.6, 4.4] g. The perfectly matched table, which redoes the computation of the targets, obtained 0.3 [0.1, 0.8] g on test dishes and 0.06 [0.05, 0.07] g on unseen ingredients. The MAE of the zero-shot models was higher than that of the USDA table, and that of the 8B higher than that of the 4B. Second attempts concerned 0.2% of zero-shot 4B answers and 1.2% of 8B answers. Energy differed by more than 25% from 4C + 4P + 9F in 46% (4B) and 75% (8B) of zero-shot answers, versus 1% after fine-tuning. On test dishes, the MAE of the fine-tuned model was 1.3 g for 1 or 2 ingredients (192 dishes), 2.7 g for 3 to 6 (114) and 3.2 g for 7 or more (106); that of the USDA table was 6.2, 13.1 and 6.8 g (Figure S1).

#### External validation: 100 real Shanghai meals

The reference carbohydrate of these meals ranged from 1.3 to 179.4 g (median 36.3 g). Against this reference, the MAE of the fine-tuned model was 12.2 [9.9, 15.2] g, versus 22.6 [18.0, 28.7] g for the USDA table; paired differences: -10.5 [-16.8, -6.0] g versus the table, -22.1 [-28.2, -16.8] g versus zero-shot 4B and -20.9 [-29.8, -15.4] g versus zero-shot 8B (Table 6 and Figure 3). The median relative error was 24% (table: 30%). Reference certainty levels were distributed as 23 certainty-1 meals, 65 certainty-2 and 12 certainty-3; restricted to certainty 2 and 3 (77 meals), the MAE was 12.0 [9.5, 15.9] g for the fine-tuned model and 23.2 [17.9, 30.6] g for the table (difference -11.2 [-18.4, -6.3] g). Line-by-line checking of the reference led to changing 0 of the 100 carbohydrate values and 0 certainty level.

**Figure 3.**
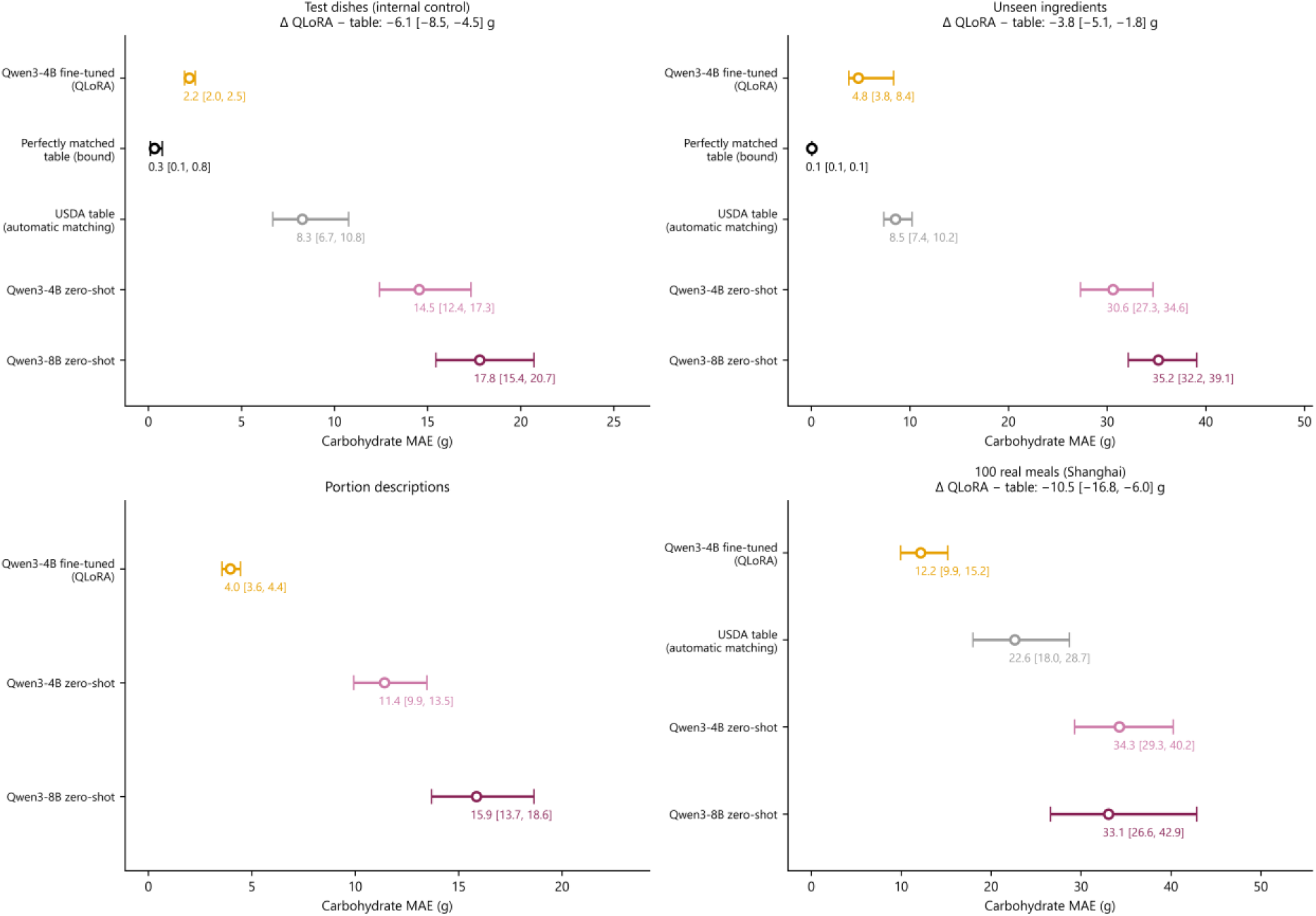
Carbohydrate MAE (circles, 95% CI) on the four sets: test dishes (internal control), unseen ingredients, portion descriptions and real Shanghai meals; perfectly matched table as a bound on Nutrition5k dishes; in each panel title, the paired difference between the fine-tuned model and the comparator.

#### Second reference, recomputed from the Chinese text

Quantities differed between the Chinese text and its English translation for 5 of the 100 meals. The second reference differed from the main reference by more than 5 g for 3 meals (maximum gap 17.7 g; median gap 0.0 g; Bland-Altman bias -0.2 g, limits -4.9 to 4.5 g). Against this second reference, the MAE of the fine-tuned model was 12.5 [10.2, 15.9] g (versus 12.2 g with the main reference) and that of the USDA table 23.0 [18.3, 29.1] g (versus 22.6 g), with a paired difference of -10.5 [-16.9, -5.9] g (versus -10.5 g): the values obtained with the two references were close.

#### Gaps by meal size

The Bland-Altman bias of the fine-tuned model on real meals was -10.3 g (constant limits of agreement -39.7 to 19.0 g). In regression, the bias varied with the mean (slope -0.52 [-0.64, -0.38]): it was -6.4 g for a 30 g meal and -32.3 g for an 80 g meal, with half-widths of the limits of agreement of 17.6 and 32.3 g; for the table, the bias slope was 0.56 [0.12, 0.87] (Figure 4).

**Figure 4.**
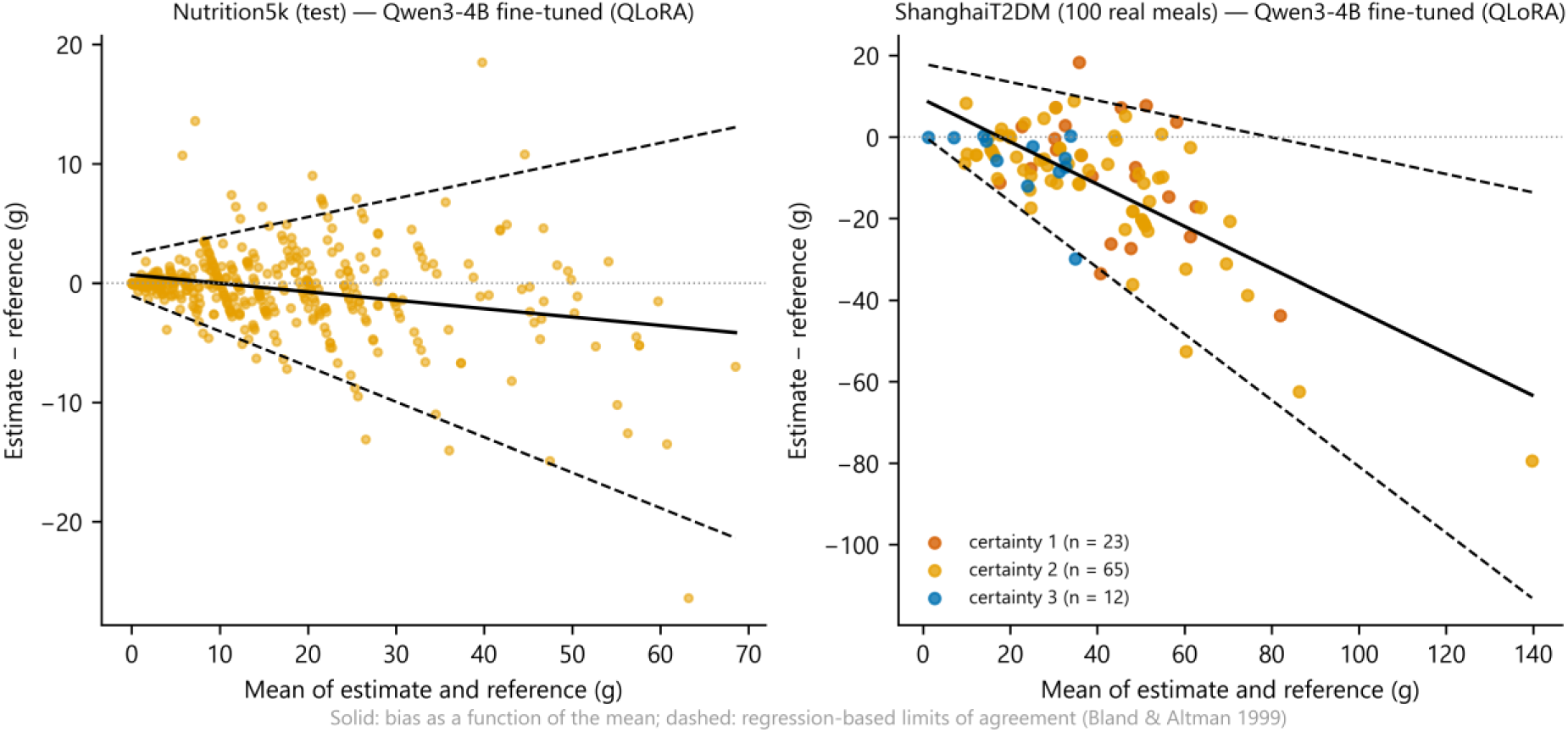
Regression-based Bland-Altman plot of the carbohydrate of the fine-tuned model: Nutrition5k (test) and real Shanghai meals, coloured by reference certainty.

The perfectly matched table (ideal bound) and the second reference of the real meals are reported in the text.

### 3.2 Step 2: prediction of the glucose rise

#### Model performance

Table 7 gives, for each model, the correlation r between predicted and observed rises and the AUROC of the peak > 180 mg/dL. M2, the personalised gradient boosting, reached r = 0.49 [0.42, 0.54] in Shanghai and 0.63 [0.55, 0.69] in CGMacros. Its correlation exceeded that of M0_spline by 0.25 [0.19, 0.32] in Shanghai and by 0.31 [0.24, 0.38] in CGMacros, and that of Chronos-2 used without training by 0.25 [0.17, 0.32] and 0.44 [0.34, 0.52]. The personal mean used alone gave r = 0.35 [0.27, 0.43] in Shanghai and 0.33 [0.26, 0.42] in CGMacros.

**Table 7.** Model performance (out-of-fold predictions, main analysis, 95% CI).

| Model | Shanghai: $r$ ($\Delta G_{\max}$) | Shanghai: AUROC (peak) | CGMacros: $r$ ($\Delta G_{\max}$) | CGMacros: AUROC (peak) |
| --- | --- | --- | --- | --- |
| Pre-meal glucose alone (level 1) | 0.38 [0.32, 0.42] | 0.80 [0.76, 0.83] | 0.45 [0.36, 0.55] | 0.77 [0.72, 0.82] |
| M0 linear (baseline + carbohydrate) | 0.23 [0.16, 0.30] | 0.79 [0.75, 0.82] | 0.10 [0.04, 0.16] | 0.78 [0.73, 0.83] |
| M0_spline (main linear reference) | 0.23 [0.17, 0.30] | 0.79 [0.75, 0.82] | 0.32 [0.25, 0.37] | 0.78 [0.73, 0.83] |
| M1 gradient boosting | 0.44 [0.37, 0.50] | 0.79 [0.74, 0.82] | 0.57 [0.47, 0.66] | 0.83 [0.79, 0.87] |
| **M2 = M1 + personalisation** | **0.49 [0.42, 0.54]** | **0.80 [0.76, 0.84]** | **0.63 [0.55, 0.69]** | **0.84 [0.80, 0.88]** |
| M3 Chronos-2 (zero-shot) | 0.23 [0.19, 0.29] | 0.80 [0.77, 0.84] | 0.20 [0.15, 0.24] | 0.78 [0.73, 0.84] |
| M2 MAE (mg/dL) | 30.6 [28.1, 32.9] | Not applicable (binary outcome) | 24.9 [22.3, 27.7] | Not applicable (binary outcome) |
| M2 $r$ for iAUC | 0.49 [0.43, 0.54] | Not applicable (continuous outcome) | 0.59 [0.47, 0.68] | Not applicable (continuous outcome) |

#### Contribution of each variable family

Families are presented, in each dataset, from the largest to the smallest contribution (difference in r from the previous level; Figures 5 and 6). In CGMacros, pre-meal glucose alone (level 1) gave r = 0.45 [0.36, 0.55]; next came carbohydrate, which increased r by 0.095 [0.068, 0.128] (Holm p 0.007), MAE by -2.52 [-3.23, -1.86] mg/dL and the peak AUROC by 0.041 [0.023, 0.064]; personalisation (0.056 [0.026, 0.093]; Holm p 0.011; MAE -1.95 [-2.90, -1.01] mg/dL); the other macronutrients (0.014 [0.001, 0.027]; Holm p 0.32); clinical variables (0.010 [-0.084, 0.077]); and finally pre-meal heart rate (-0.003 [-0.030, 0.012]). In Shanghai, pre-meal glucose alone gave r = 0.38 [0.32, 0.42]; next came personalisation (0.048 [0.017, 0.083]; Holm p 0.055), clinical and treatment variables (0.036 [-0.020, 0.083]), the other macronutrients (0.022 [-0.002, 0.049]; Holm p 0.76) and carbohydrate (0.004 [-0.010, 0.017]; MAE -0.15 [-0.47, 0.18] mg/dL).

**Figure 5.**
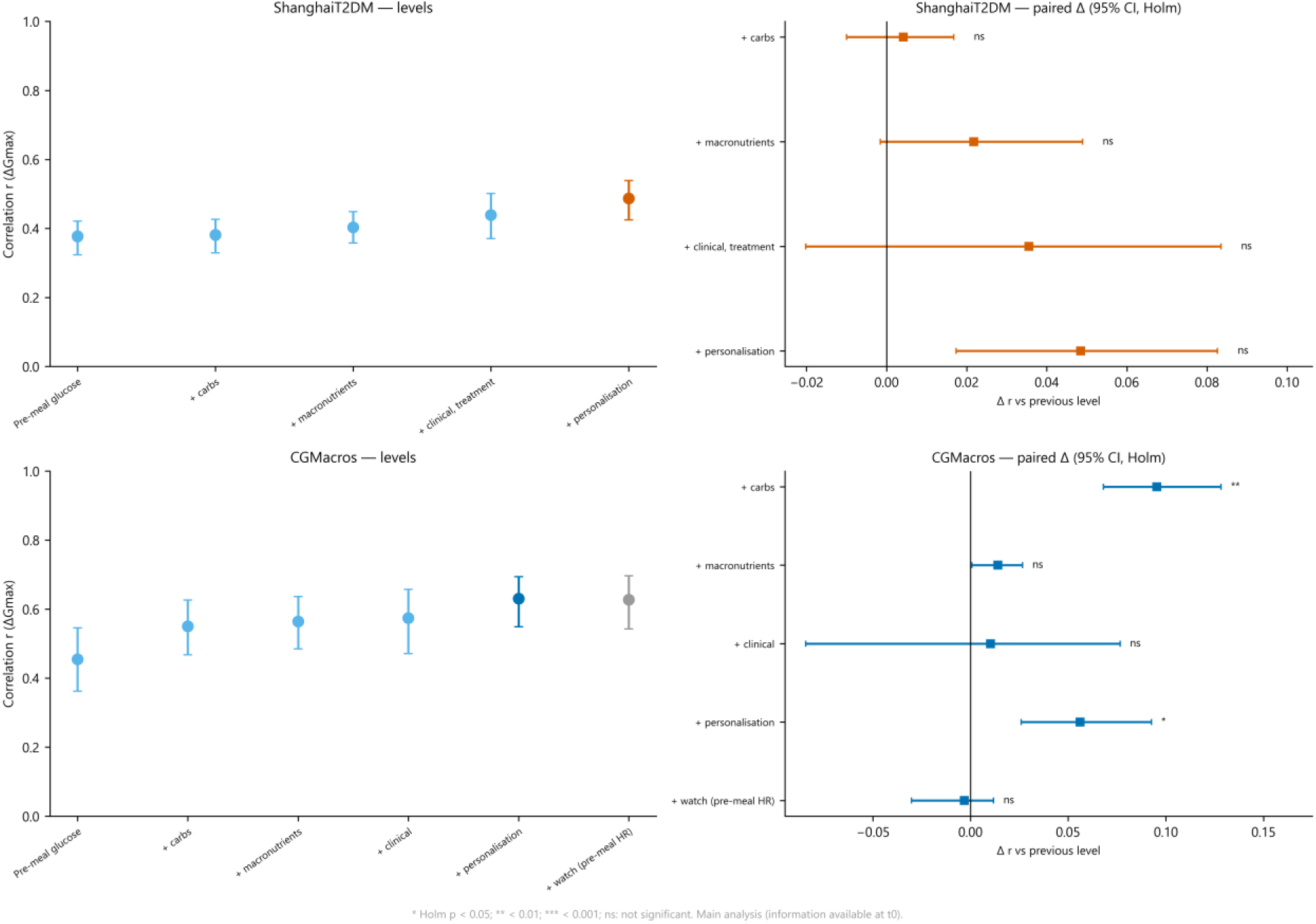
Correlation r between predicted and observed rises (ΔGmax) as variable families are added: successive levels (points and 95% CI, left) and paired difference from the previous level (95% CI, stars: Holm p, right); top ShanghaiT2DM, bottom CGMacros. Main analysis (information available at t0).

**Figure 6.**
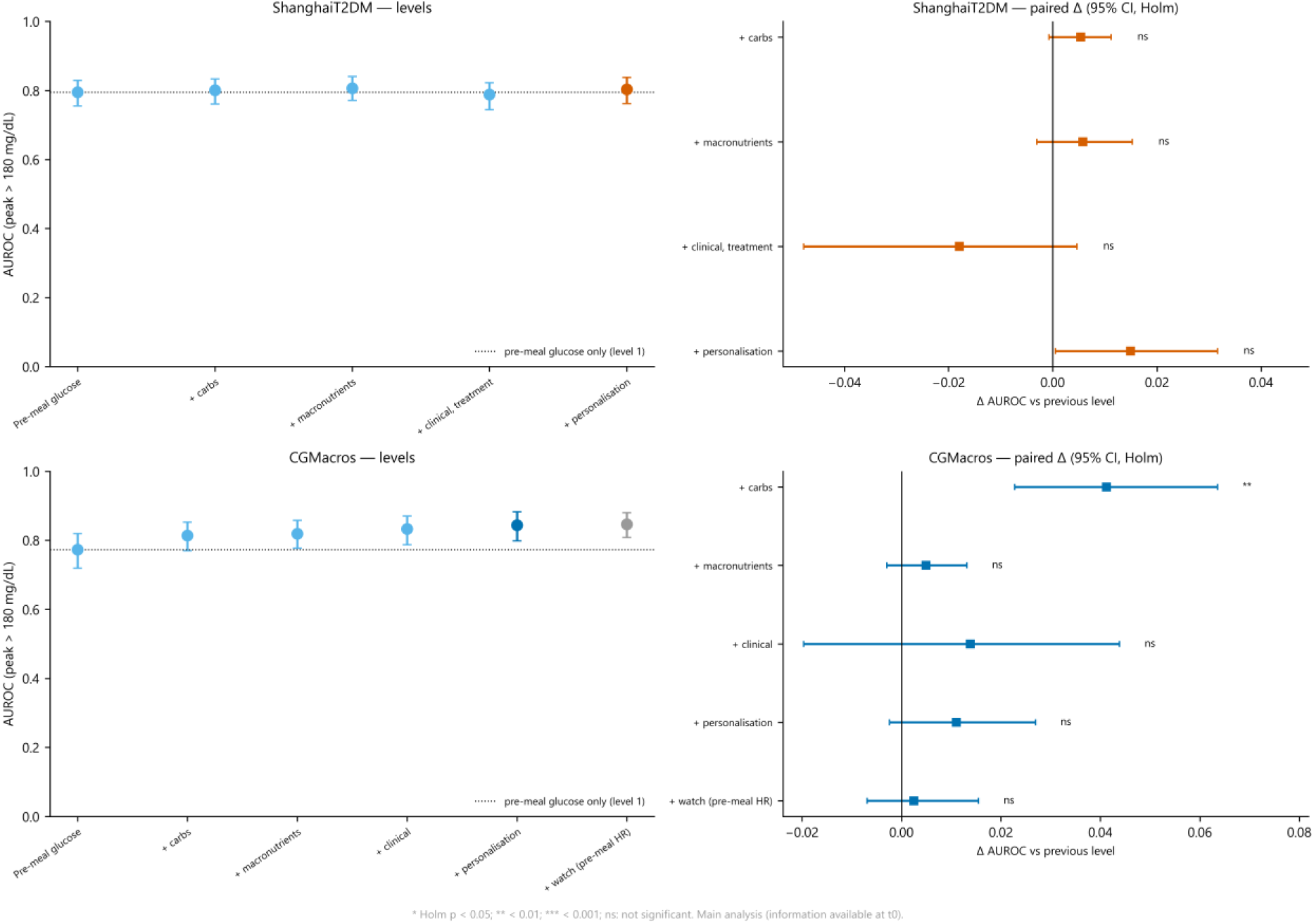
AUROC of the peak > 180 mg/dL as variable families are added: successive levels (dotted line: pre-meal glucose alone) and paired difference from the previous level; top ShanghaiT2DM, bottom CGMacros.

#### Contribution of carbohydrate: comparison between the two datasets

The r gain from carbohydrate was larger in CGMacros: difference between gains (CGMacros - Shanghai) of 0.091 [0.060, 0.126] (p < 0.001), in the same direction for MAE (-2.37 [-3.11, -1.62] mg/dL) and R² (0.105 [0.070, 0.141]). In the harmonised analysis (same level-1 variables in both datasets, meals with at least 15 g), the gain remained positive in the 14 CGMacros people with T2D (0.050 [0.017, 0.088]; 361 meals); in Shanghai, its CI included 0 in both subsets studied: meals eaten without prandial insulin, AGI or secretagogue (-0.007 [-0.028, 0.011]; 1,027 meals, 63 people) and people without insulin or secretagogue (0.008 [-0.027, 0.040]; 854 meals, 37 people). The gap with the CGMacros people with T2D was 0.057 [0.018, 0.102] and 0.041 [-0.005, 0.096], respectively. On the 100 annotated meals, the Spearman correlation between ΔGmax and carbohydrate was 0.175 [-0.003, 0.374] with the reference and 0.113 [-0.088, 0.291] with the LLM; the slope of ΔGmax per 10 g of carbohydrate was 3.1 [-1.0, 7.5] and 3.4 [-2.1, 9.7] mg/dL (LLM / reference ratio: 1.10 [-0.99, 5.81]).

#### Nutrient source (Shanghai)

With M2 retrained on each nutrient source, r was 0.49 with the fine-tuned LLM, 0.49 with zero-shot 4B, 0.50 with zero-shot 8B, 0.49 with the USDA table and 0.49 with the raw sum of grams; difference fine-tuned LLM - sum of grams: -0.006 [-0.023, 0.008].

#### Error, calibration and secondary outcomes

The MAE of M2 was 30.6 [28.1, 32.9] mg/dL in Shanghai and 24.9 [22.3, 27.7] mg/dL in CGMacros (RMSE 39.0 and 33.5; out-of-fold R² 0.22 and 0.39). Its skill score was 0.15 and 0.25 relative to the population mean, and 0.07 [-0.01, 0.12] and 0.19 [0.09, 0.25] relative to the previous personal mean. The within-person correlation of M2 predictions was 0.50 [0.46, 0.54] in Shanghai (78 people) and 0.59 [0.53, 0.63] in CGMacros (44 people); that of glucose alone (level 1) was 0.44 [0.39, 0.49] and 0.34 [0.26, 0.42]. For the peak AUROC, the difference M2 - M0_spline was 0.019 [-0.016, 0.049] in Shanghai and 0.061 [0.027, 0.093] in CGMacros; M2 - M0: 0.018 [-0.015, 0.048] and 0.067 [0.029, 0.102]. For a rise above 50 mg/dL, the AUROC of M2 was 0.74 [0.71, 0.77] in Shanghai and 0.80 [0.77, 0.83] in CGMacros, versus 0.68 [0.65, 0.71] and 0.64 [0.58, 0.69] for glucose alone and 0.61 [0.57, 0.65] and 0.61 [0.58, 0.64] for M0_spline (thresholds of 30 and 70 mg/dL in Shanghai: 0.77 and 0.73). For meals with starting glucose below 140 mg/dL, the AUROC of M2 for the peak > 180 mg/dL was 0.71 [0.65, 0.76] in Shanghai (1,465 meals) and 0.79 [0.74, 0.84] in CGMacros (955 meals), versus 0.69 and 0.66 for glucose alone. The calibration slope of M2 for ΔGmax was 0.77 (Shanghai) and 0.92 (CGMacros); for the peak, 0.68 and 0.59 (Figures S2 and S3).

#### Subgroups

In Shanghai, r of M2 was 0.50 [0.41, 0.58] for meals on insulin (1,010 meals), 0.44 [0.25, 0.59] on sulfonylurea or glinide (373) and 0.52 [0.46, 0.59] on neither (956); in CGMacros, 0.66 [0.56, 0.74] in people with T2D, 0.53 [0.37, 0.65] in people with prediabetes and 0.54 [0.47, 0.61] in normoglycaemic people. By sex, r was 0.46 [0.40, 0.52] in women and 0.45 [0.36, 0.53] in men in Shanghai, 0.66 [0.56, 0.74] and 0.57 [0.45, 0.66] in CGMacros; in CGMacros, 0.63 [0.54, 0.70] in people of Hispanic or Latino origin and 0.59 [0.51, 0.68] in others.

#### Leave-one-family-out ablation, Shanghai sensitivity analyses and personalisation

Removing nutrients from M2 changed r by 0.005 [-0.018, 0.023] in Shanghai and by 0.121 [0.082, 0.166] in CGMacros (M2 minus M2 without the family; Holm p 1.000 and 0.007); removing pre-meal glucose, by 0.061 [0.014, 0.098] and 0.073 [0.038, 0.107]; removing personalisation, by 0.048 [0.017, 0.083] and 0.056 [0.026, 0.093]; removing treatment (Shanghai), by 0.034 [0.004, 0.055]. In Shanghai, without the 6 recordings whose “intravenous insulin” column was filled, r of M2 was 0.49 [0.42, 0.54] and the carbohydrate gain 0.002 [-0.017, 0.017]; without the 35 pump recordings, 0.48 [0.41, 0.55] and 0.010 [-0.005, 0.027]; without the prandial insulin dose as a predictor, r of M2 was 0.48 [0.42, 0.53]. With meals of at least 15 g of carbohydrate (1,994 meals), the carbohydrate gain in Shanghai was -0.004 [-0.018, 0.008]. The k estimated by the method of moments was 4.3 in Shanghai and 4.6 in CGMacros; with k = 1, 3, 10 and estimated k, r of M2 was 0.48, 0.49, 0.49 and 0.49 in Shanghai, and 0.63, 0.63, 0.63 and 0.63 in CGMacros.

#### Diagnostics of the linear references

In CGMacros, M0 (baseline glucose and carbohydrate, linear) gave r = 0.10 [0.04, 0.16], versus 0.32 [0.25, 0.37] for M0_spline. The relationship between baseline glucose and the rise changed direction depending on the level of analysis: correlation of -0.02 over all meals, 0.62 between people’s means and -0.33 within people (Shanghai: -0.25, 0.03 and -0.40). Entered CGMacros carbohydrate reached 761 g. In M0, the mean standardised coefficients were -0.57 for baseline glucose and 6.98 for carbohydrate; the per-fold correlation ranged from 0.09 to 0.26, and 0 values had been imputed. With glucose alone, r was -0.13 [-0.22, -0.02] with a linear model and 0.07 [0.02, 0.13] with gradient boosting; a linear model on the level-1 variables gave 0.25 [0.17, 0.34] and gradient boosting with glucose and carbohydrate 0.49 [0.42, 0.57]. In Shanghai, r was 0.23 [0.17, 0.30] for M0_spline and 0.15 [0.08, 0.22] for gradient boosting with glucose alone (Figure S5).

#### Stability and sample size

Over five repeated splits, r of M2 ranged from 0.51 to 0.55 in Shanghai (standard deviation 0.016) and from 0.60 to 0.65 in CGMacros (standard deviation 0.016); the carbohydrate gain ranged from 0.000 to 0.011 and from 0.087 to 0.098. With the observed out-of-fold R² (0.22 and 0.39) and 31 and 18 candidate variables, the minimum required sample size was 1,131 meals in Shanghai and 312 in CGMacros, for 2,339 and 1,286 available meals from 99 and 45 people. Compared with the sensitivity analysis with information up to +30 min and +2 h, r of M2 went from 0.47 to 0.49 in Shanghai and from 0.62 to 0.63 in CGMacros (Table S2).

### 3.3 Step 3: transfer between datasets

Trained in CGMacros (T2D and prediabetes) and applied to ShanghaiT2DM (A1), the model had a correlation of 0.37, versus 0.48 in internal validation on the same meals (Δr = -0.11 [-0.17, -0.05]) and 0.49 for full M2; the peak AUROC was 0.80 versus 0.78 (Δ = 0.023 [-0.007, 0.072]). Trained in the whole CGMacros dataset (A2), Δr = -0.11 [-0.16, -0.05]. Trained in Shanghai and applied to the whole CGMacros dataset (B2), it had a correlation of 0.55 versus 0.67 (Δr = -0.12 [-0.19, -0.04]) and an AUROC of 0.80 versus 0.86 (Δ = -0.054 [-0.088, -0.016]); towards the CGMacros people with T2D (B1), Δr = -0.03 [-0.11, 0.07]. With the Libre Pro, Δr = -0.09 [-0.14, -0.02] (A1L) and 0.00 [-0.08, 0.16] (B1L).

Before and after recalibration, the calibration slope was 0.46 [0.37, 0.58] and 0.68 [0.56, 0.86] (A1), 0.52 [0.41, 0.65] and 0.68 [0.53, 0.84] (A2), 1.28 [0.99, 1.59] and 0.74 [0.57, 0.92] (B1), 0.99 [0.83, 1.20] and 0.82 [0.69, 0.99] (B2), 0.59 [0.50, 0.73] and 0.69 [0.58, 0.85] (A1L), 1.34 [1.10, 1.59] and 0.85 [0.71, 1.02] (B1L). The recalibration slope learned on the 20% of people and the slope that would have been optimal on the remaining 80% were 1.74 and 1.28 for B1 (2 recalibration people), and 0.67 and 0.46 for A1. The MAE of raw, recalibrated and re-estimated transfer was 38.7, 32.8 and 32.1 mg/dL for A1, and 27.4, 30.5 and 25.5 mg/dL for B2; after re-estimation, r was 0.44 (A1) and 0.62 (B2) (Figure 7).

**Figure 7.**
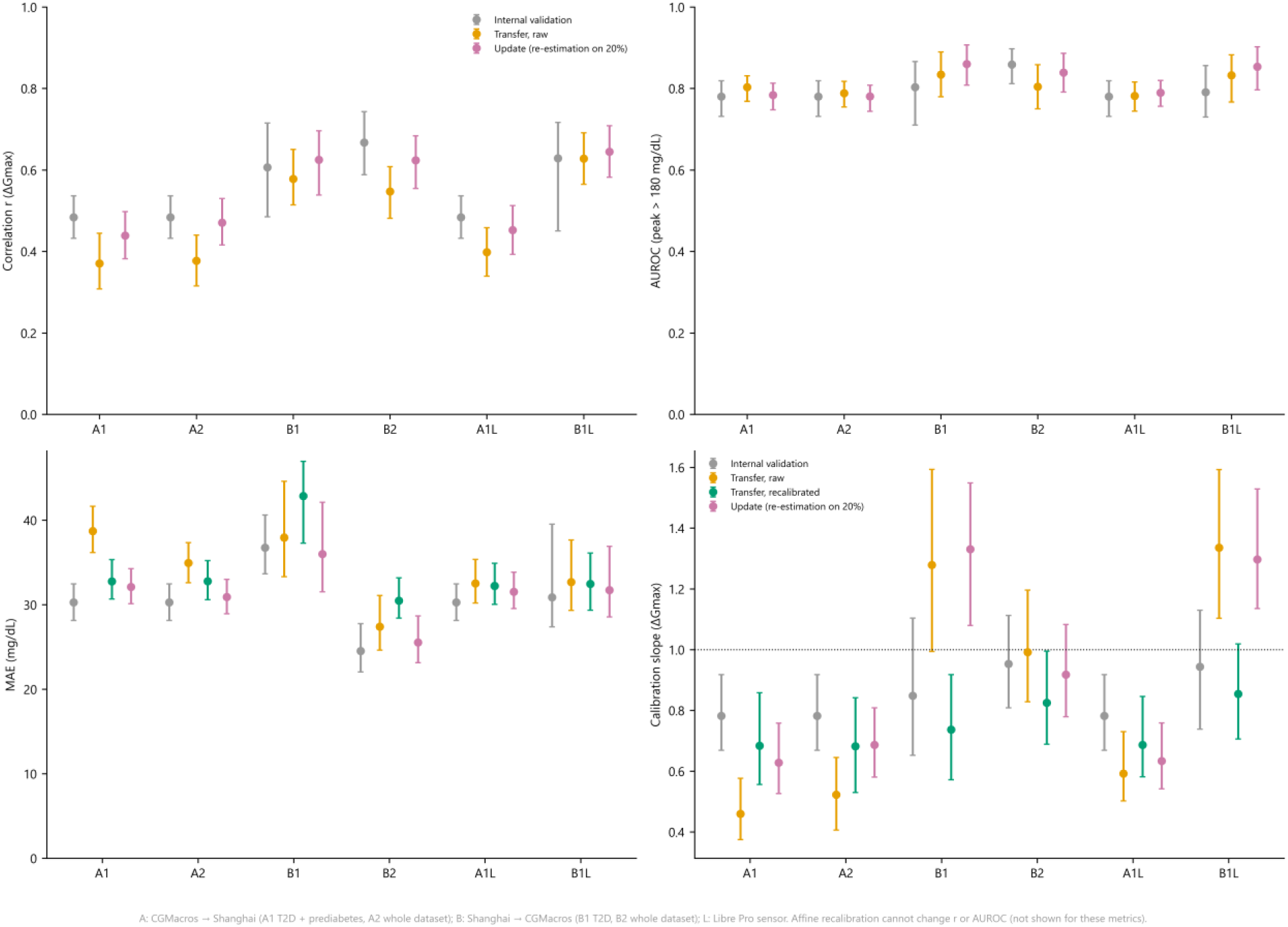
Transfer between datasets, six scenarios: correlation and AUROC (top; internal validation, raw transfer, update by re-estimation), MAE and calibration slope (bottom; with recalibration), points and 95% CI. A: CGMacros → ShanghaiT2DM; B: ShanghaiT2DM → CGMacros; L: Libre Pro sensor.

### 3.4 Complementary analyses

#### Carbohydrate variability and relationship with the rise

The coefficient of variation of carbohydrate per meal was 0.50 in Shanghai and 0.64 in CGMacros. The within-person correlation between carbohydrate and the rise was 0.081 [-0.005, 0.152] in Shanghai (78 people), 0.300 [0.254, 0.347] in CGMacros and 0.278 [0.174, 0.367] in the CGMacros people with T2D.

#### Treatments taken at mealtime (Shanghai)

Prandial insulin accompanied 24% of Shanghai meals and an AGI 26%. For the 1,387 meals eaten with neither, the within-person correlation between carbohydrate and the rise was 0.220 [0.144, 0.295] (paired difference with all meals, in the 48 people with at least 8 meals in both sets: 0.000 [-0.046, 0.044]) and the r gain from carbohydrate -0.003 [-0.025, 0.021]; in the CGMacros people with T2D, this gain was 0.050 [0.017, 0.088].

#### Links between carbohydrate and other predictors

Carbohydrate was correlated with starting glucose (ρ = -0.07 in Shanghai, -0.06 in CGMacros), time (-0.17 and -0.10), meal rank (-0.18 and -0.11) and, in Shanghai, prandial insulin dose (0.10).

#### Food categories

Adding grams per food category to M2 (Shanghai) changed r by 0.008 [-0.009, 0.027] (uncorrected p 0.443; Holm p 1.00).

#### Variable importance

In grouped permutation, the family whose shuffling increased M2 MAE the most was, in Shanghai, pre-meal glucose (3.7 mg/dL), ahead of time and rank (2.1), personalisation (1.4) and nutrients (0.8); in CGMacros, nutrients (4.8 mg/dL), ahead of pre-meal glucose (4.2), clinical variables (4.1) and personalisation (3.0) (Figure 8).

**Figure 8.**
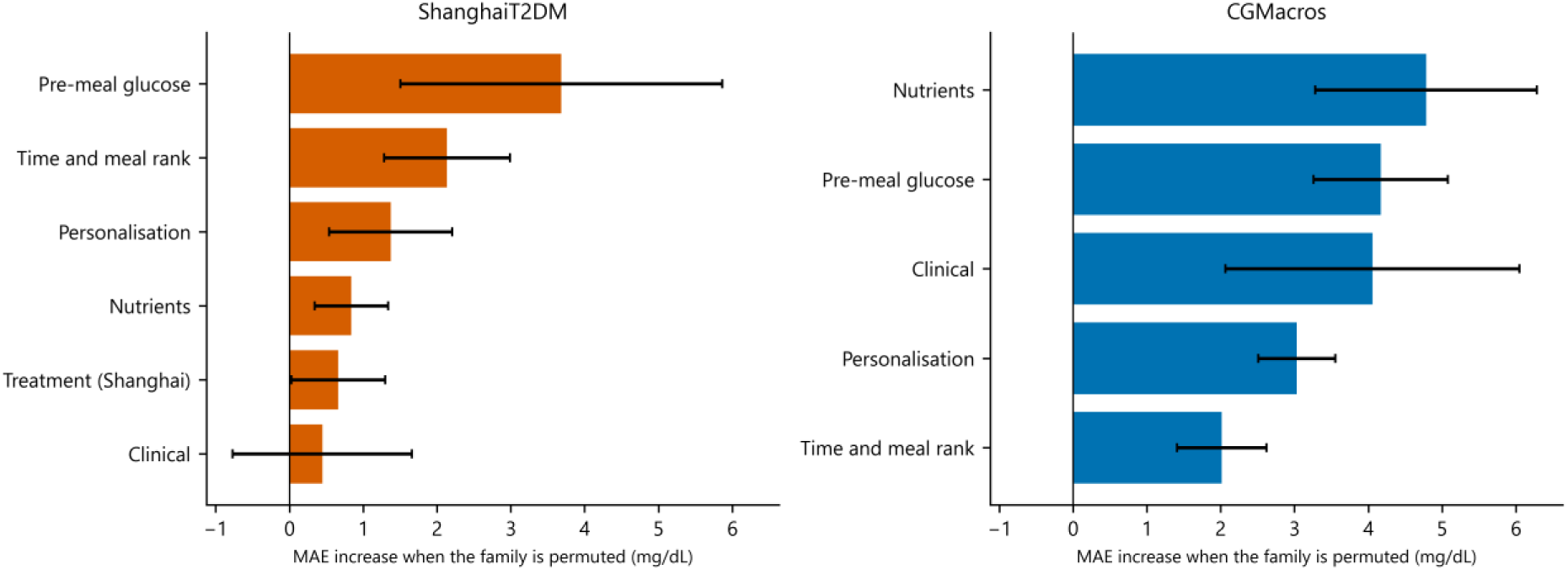
Importance of M2 variable families by grouped permutation (mean and standard deviation over the 5 folds; same scale for both datasets).

#### Clinical utility of the alert

At a probability threshold of 0.5, the net benefit of the M2 “peak > 180 mg/dL” alert was 0.21 [0.13, 0.28] in Shanghai and 0.16 [0.09, 0.24] in CGMacros; ΔNB (M2 - M0_spline): 0.018 [-0.019, 0.051] and 0.034 [0.010, 0.060]. Over thresholds 0.2 to 0.8, the CI of ΔNB excluded 0 for 0 of the 13 thresholds in Shanghai and for 6 in CGMacros; ΔNB ranged from -0.010 to 0.020 in Shanghai and from 0.011 to 0.051 in CGMacros (Figure 9).

**Figure 9.**
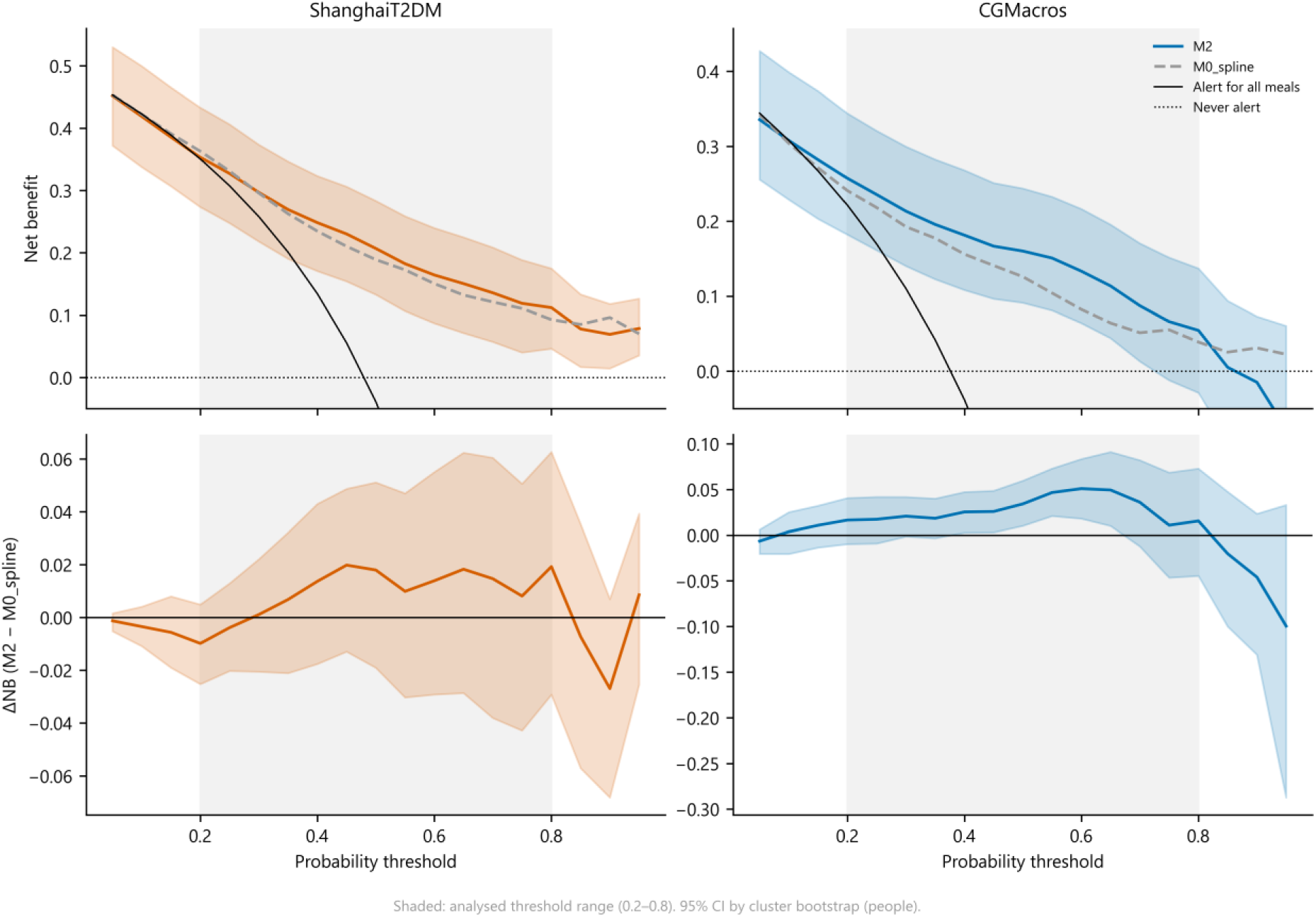
Decision curves of the “peak > 180 mg/dL” alert (net benefit of M2 with 95% CI, M0_spline, alert for all) and net-benefit difference M2 - M0_spline; shaded area: thresholds 0.2 to 0.8.

#### Curves by carbohydrate tertile

Carbohydrate tertiles ranged from 1 to 24 g, 24 to 40 g and 40 to 142 g in Shanghai, and from 15 to 40 g, 40 to 66 g and 66 to 761 g in CGMacros. At 60 minutes, the mean rise of tertiles 1, 2 and 3 was 24 [20, 30], 33 [27, 39] and 36 [29, 43] mg/dL in Shanghai, and 19 [14, 23], 40 [31, 49] and 36 [29, 43] mg/dL in CGMacros; mean ΔGmax: 45, 54 and 56 mg/dL in Shanghai, 35, 60 and 56 mg/dL in CGMacros (Figure 10).

**Figure 10.**
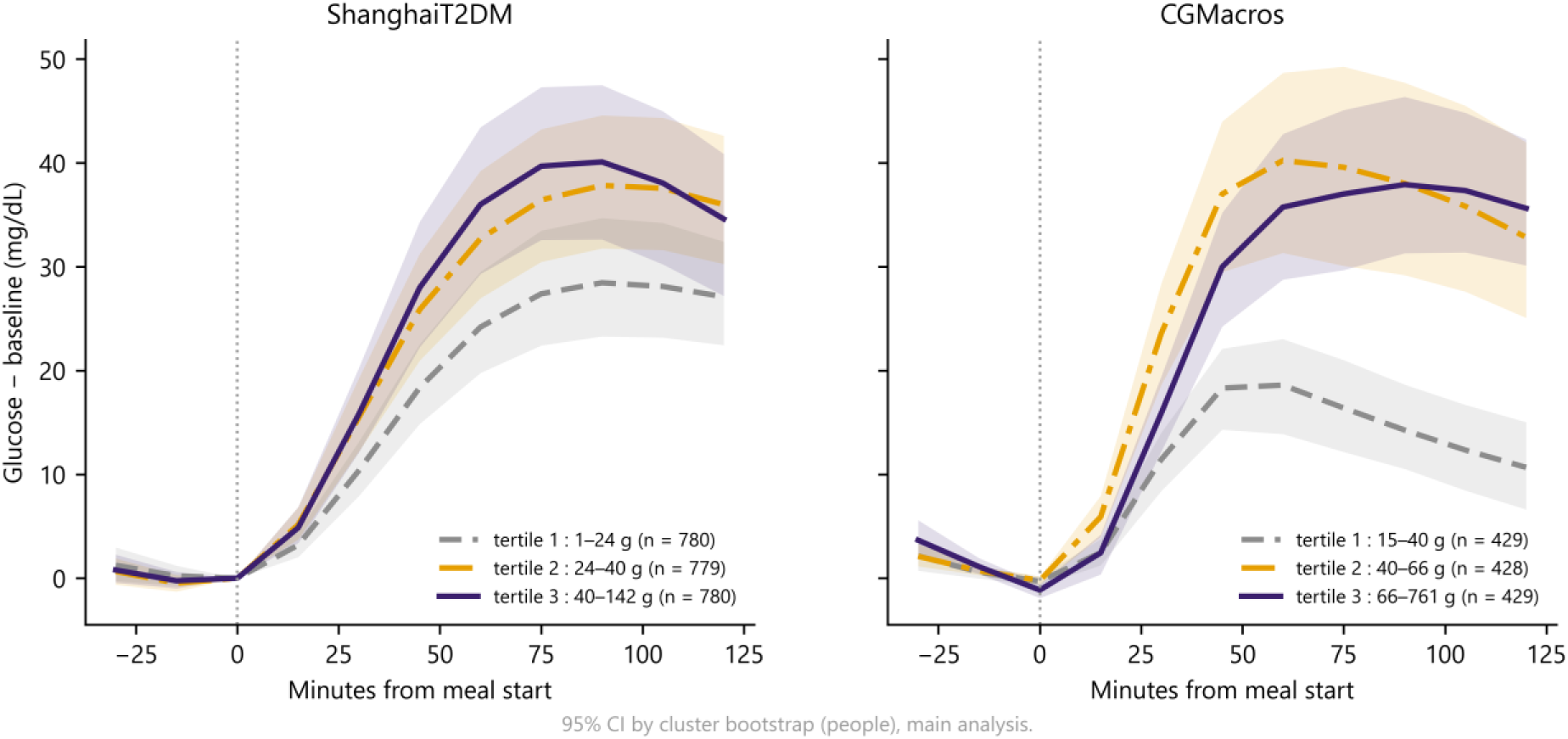
Mean post-meal glucose curve by carbohydrate tertile (bounds in the legend; 95% CI by cluster bootstrap).

### 3.5 Sensitivity analyses and sensor agreement

With an exclusion window of 60 or 120 minutes instead of 90 (analysis with information up to +2 h), r was 0.46 and 0.45 in Shanghai (same analysis with 90 minutes: 0.47), and 0.63 and 0.62 in CGMacros (0.62). With the Libre Pro as the CGMacros sensor, r was 0.67 [0.59, 0.73]. Recomputed on the native Dexcom measurements (1,286 meals), median ΔGmax was 47 mg/dL, versus 41 mg/dL on the 15-minute grid (median gap 4.1 mg/dL; ρ = 0.98); r of M2 for this rise was 0.63 [0.54, 0.69].

In the same CGMacros people, the Libre Pro recorded 606 episodes below 70 mg/dL for at least 15 minutes, versus 48 for the Dexcom. Of the Libre’s low minutes, 1.5% were also low for the Dexcom, whose median value at those times was 107 mg/dL (Figure S4). In Shanghai, of 186 low sensor readings with a capillary glucose within ±15 minutes, 24 were found by capillary glucose.

## 4. Discussion

### 4.1 Main findings

Two positive results stand out. The first concerns carbohydrate counting: an open 4-billion-parameter LLM, fine-tuned with QLoRA on public data, estimates the carbohydrate of real patient meals with an error almost half that of an automatically matched USDA table, and nearly a third of that of the same model without fine-tuning. This gain is found on the meals with the most certain reference, with the reference recomputed from the Chinese text, and on dishes containing ingredients never seen in training: the model has therefore not merely memorised the Nutrition5k dishes. The second concerns glucose: personalised gradient boosting (M2) roughly doubles the correlation of the linear reference in both cohorts, outperforms the Chronos-2 foundation model used without training and, for a rise above 50 mg/dL, an outcome independent of the starting level, does better than pre-meal glucose alone in both datasets. With r = 0.63 in CGMacros, M2 is in the range of published personalised models (r = 0.62 to 0.69 [12, 21]), without microbiome data or standardised test meals.

These results need context. The error on real meals is several times that on test dishes, and the model underestimates more as the meal grows larger. On Nutrition5k dishes, the perfectly matched table does better than the model; this is not a failure of the LLM but a property of the measurement: the Nutrition5k target carbohydrate was computed with these same contents, so this table redoes the computation of the target and can hardly be wrong. It is available only when each ingredient is identified without ambiguity, which is never the case for a patient’s unstructured text; on real meals, the table that can actually be used, automatically matched, makes almost twice the error of the LLM. In step 2, the contribution of carbohydrate differs between datasets: nil in ShanghaiT2DM, where no nutrient source did better than a plain sum of grams, positive in CGMacros, with a between-dataset difference whose CI is reported. In Shanghai, the peak > 180 mg/dL AUROC of the full model remains close to that of pre-meal glucose alone, and in CGMacros the gap remains modest: this outcome, dominated by the starting level, poorly captures the model’s contribution, hence the value of outcomes based on the rise.

### 4.2 A confounded between-dataset comparison, and a chain tested in a single dataset

The gap between the two datasets cannot be attributed to a single cause. ShanghaiT2DM and CGMacros differ simultaneously in population (T2D only versus a minority with T2D, mostly of Hispanic or Latino origin), glycaemic control, treatments, care setting, sensor (Libre H at 15 minutes versus Dexcom G6), meal size and variability, and above all the source of carbohydrate: estimated by the LLM, with underestimation, in Shanghai; entered in the dataset in CGMacros. The harmonised analysis reduces these differences without removing them: even restricted to meals eaten without prandial insulin, AGI or secretagogue, or to people without insulin or secretagogue, the carbohydrate gain remained small in Shanghai, which argues against an explanation by mealtime treatment alone. On the 100 annotated meals, the relationship between carbohydrate and the rise was weak with the reference as with the LLM, and the CI of the slope ratio was too wide to quantify the attenuation due to LLM error. Finally, as CGMacros contains no textual meal description, the full “text → LLM → glucose” chain was evaluated in ShanghaiT2DM only, that is, where carbohydrate added nothing: the positive CGMacros result concerns entered carbohydrate, not the LLM. The CGMacros meal photographs would allow this chain to be tested with a vision model, which would be another study.

### 4.3 Comparison with the literature

Zagaroli et al. compared four commercial LLMs with a reference computed by clinicians on meals described in text: only one reached equivalence within ±5% [18]. The meals, reference and metrics differ from ours, which precludes a direct numerical comparison; our work shows that an open 4-billion-parameter model can be fine-tuned for the same task and used locally. The weakness of carbohydrate for predicting the glycaemic response in Shanghai is consistent with Shen et al., who obtained very low correlations with carbohydrate alone in the same dataset [21]. Their higher overall correlations partly stem from a meal-level split, in which the same people appear in training and testing; with a person-level split, they remain higher than ours (R = 0.69 for the maximum rise), with different meal definitions and variables, notably finer food categories from a nutrition database. Our person-level validation measures what the model would be worth in a new patient. In adults without diabetes, a personalised model outperformed carbohydrate alone (R = 0.62 versus 0.40) [12]; our correlations remain below those obtained with richer data, notably the gut microbiome [11, 13].

### 4.4 Interpretation

Several mechanisms, not mutually exclusive and not tested causally, may contribute to the weakness of carbohydrate in Shanghai: co-interventions at mealtime (prandial insulin is dosed according to the meal and AGIs are taken with it), a more closely monitored care setting (nearly a third of recordings with an insulin pump; weighed, smaller and less variable meals), the nature of carbohydrate (glycaemic index, food matrix), which quantity does not capture [28, 55], and measurement error in carbohydrate itself. In Shanghai, the relationship between carbohydrate and the rise was weak, overall as within people, even though the richest meals rose more on average (tertiles); carbohydrate was moreover only weakly correlated with time, meal rank and starting glucose, already present at level 1, so that such a small effect cannot be distinguished from noise once these variables are known. The carbohydrate gain in the 14 CGMacros people with T2D suggests that diabetes itself does not suffice to explain the gap. The 15-minute resolution and the sensors’ own imprecision add noise [56].

Linear M0 performed poorly in CGMacros because the relationship between starting glucose and the rise is weak there and of different sign between and within people, and because extreme entered carbohydrate values weigh on a linear model; no implementation error was found. This is why the main linear reference is M0_spline, which allows a curved relationship. Calibration slopes below 1 mean that M2 predictions are too spread out and should be recalibrated before any individual use. An affine recalibration does not change the ranking of meals, hence neither r nor the AUROC; it sometimes worsened the slope in the target dataset because it was estimated on only 20% of the people, with a marked person effect: the slope learned on these few people did not hold for the others. Finally, the strong sensor disagreement in the low range justifies not having conducted any hypoglycaemia analysis on these datasets.

#### Two models rather than one

The same LLM could have been fine-tuned a second time to predict the glucose rise directly. This option was ruled out at the design stage, for four reasons. (1) Nature of the data: once the meal is converted into nutrients, all step-2 inputs are numbers (glucose values, time, doses, clinical characteristics, the person’s history); on such tabular data, gradient-boosted decision trees perform at least as well as deep neural networks and remain the reference method [57, 58]. (2) Size: a few thousand meals from about a hundred people per dataset are enough for gradient boosting, but too few to tune without overfitting a 4-billion-parameter model whose output is text, not a calibrated continuous value. (3) Readability: separating the two tasks makes it possible to measure the error specific to each step, to swap the nutrient source with the model held constant (fine-tuned or not, table, plain sum of grams) and to estimate the weight of each variable family. (4) Use: gradient boosting trains and runs in seconds on an ordinary computer and is easily retrained in a new population. The results support this choice. M2 clearly outperformed Chronos-2, a neural network pre-trained on time series and run locally here, and all its information came from numerical variables (pre-meal glucose, nutrients, the person’s usual response), which a tree model uses directly. Separating the two tasks also made it possible to establish that, in Shanghai, counting carbohydrate more accurately did not change the prediction of the rise: a single model trained end to end from text to glucose would have masked this result. Such a single model could be justified with much larger data, but current open datasets, of about a hundred people each, are too small to test it.

### 4.5 Strengths and limitations

Strengths: protocol, splits and pass criterion set before training; evaluation of the LLM on held-out sets increasingly distant from training (test dishes, new combinations, unseen ingredients, portion descriptions, real meals translated from Chinese); reference for real meals computed by an explicit program, checked line by line and recomputed from the original text; comparison of nutrient sources with the model held constant, down to a plain sum of grams; explicit eligibility criteria at each level; defined time of prediction and main analysis restricted to information available at t0; person-level validation; two datasets and an explicit transfer; clustered CIs, paired differences and correction for multiple comparisons; calibration, error measures and clinical utility reported with their CIs; reproducibility (open data frozen by hash, fixed seed, automatically generated numbers); model executable locally, without transmission of patient data.

Limitations: (1) Nutrition5k targets are computed from masses and a table; on these dishes, a perfectly matched table does better than the model, and the main step-1 result is that of real meals. (2) The reference of the 100 meals relies on the USDA table, which does not contain all Chinese foods (mantou is approximated by white bread, for example), and on standard recipes; it was checked by a single investigator, not blinded to the hypothesis, and has not been compared with an independent annotation by a dietitian. (3) The “table” comparator relies on automatic name matching: it represents software without AI, not the work of a dietitian; a matching sheet of the Nutrition5k ingredients is provided for human checking. (4) Fine-tuning was performed once, with a single seed and an effective batch of 64 instead of the planned 32; run-to-run variability, a few-shot zero-shot comparator and possible memorisation of Nutrition5k by the base model were not evaluated. (5) The full chain was evaluated in ShanghaiT2DM only; CGMacros nutrients are estimates entered in the dataset. (6) The between-dataset comparison is confounded by many differences in population, care, sensor and source of carbohydrate. (7) Sample sizes remain modest (45 people in CGMacros, of whom 14 with T2D, a single centre, a mostly Hispanic or Latino population) and, although the number of meals exceeds the computed minimum sample size (1,131 and 312 meals), this calculation ignores the clustering of meals within people, so precision depends mainly on the number of people; the CGMacros people with T2D were not treated with insulin or a secretagogue. (8) Shanghai patients were recruited in hospital and some were on intensive insulin therapy; the inpatient or outpatient status of each recording is not reported, and the recording period covers the COVID-19 pandemic. (9) 22% of Shanghai meals had no description. (10) The Shanghai texts, translated by the dataset authors, were not evaluated in Chinese. (11) No person with type 1 diabetes was analysed, by design. (12) The protocol was not publicly deposited before the analyses; complementary and sensitivity analyses are exploratory (Table S1). (13) Patients and the public were not involved in the design.

### 4.6 Contribution of the study and role of the clinician

This work offers three lessons. First, an open, local and measurable “text → carbohydrate” conversion tool exists, with a known error on real meals and a tendency to underestimate large meals. Second, in ShanghaiT2DM, the priority for predicting glucose was not counting carbohydrate more accurately but knowing pre-meal glucose and the patient’s usual response; whether this holds for other treated people with T2D remains to be established. Third, a model trained in one dataset does not apply as is to another, which requires a local, Tunisian validation before any use.

The physician played a role here that the tool does not replace: asking the clinical question and choosing outcomes meaningful to the patient; defining what a meal, a snack and a treatment “with the meal” are, and designating prandial insulin and AGIs as confounders to be isolated; checking food by food the reference of the 100 real meals, from the Chinese text; recognising that the Libre sensor hypoglycaemias were artefacts and ruling out any hypoglycaemia analysis. The whole analysis chain is written in Python, from meal selection to the computation of the reference and the statistics; each step was reviewed and its results checked by the investigator, and each number in the manuscript is automatically linked to its source.

### 4.7 Implications and perspectives

For the patient, a local assistant able to turn “a plate of couscous and two pieces of bread” into grams of carbohydrate is conceivable and respectful of privacy, as the model can run without sending data to a server; its use on a phone was not evaluated. A monitoring application coupled with a glucose sensor is described as a perspective, not evaluated (Figure S6): time-stamped meals and treatments, conversion of the meal into nutrients, progressive personalisation and, beyond a data threshold to be defined, adaptation to the patient by individual fine-tuning, an approach that has improved glucose prediction in people with T2D [59]. Such a tool would first be a decision aid; support for calculating the prandial insulin dose could be studied at a later stage, after specific validation, and would fall under the regulation of software as a medical device.

Next steps: (1) have the real meals annotated independently by a dietitian blinded to the model outputs and to the current reference, from the Chinese text and a Chinese food composition table, then measure the agreement between the two references (intraclass correlation coefficient and Bland-Altman); (2) repeat fine-tuning with several seeds and the planned batch size, compare it with a few-shot model and with a split of dishes by ingredient combination, and look for memorisation of Nutrition5k by the base model; (3) adapt the model to Maghrebi cuisine and the Tunisian dialect, with a local food composition table; (4) validate in free-living Tunisian people with T2D wearing a sensor, with time-stamped meals; (5) assess clinical benefit in a controlled trial, along the lines of personalised nutrition trials [14, 15].

## 5. Conclusion

QLoRA fine-tuning of a small open LLM, executable offline, yields a carbohydrate estimator whose error on real patient meals is almost half that of an automatically matched table, with an underestimation of large meals. Personalised gradient boosting, trained on pre-meal glucose and the person’s usual response, doubles the correlation of a linear reference for predicting the glucose rise in two cohorts. In ShanghaiT2DM, the prediction of the glucose rise relied first on pre-meal glucose and individual history, and carbohydrate added nothing; in CGMacros, it improved it. This gap, observed between two datasets that differ in many respects, calls for validation in a population where the full chain can be tested.

## Supporting information

Supplementary_TRIPOD-AI_checklist

## Data Availability

All data used are publicly available: ShanghaiT2DM (figshare, CC BY 4.0; doi:10.1038/s41597-023-01940-7), CGMacros (PhysioNet; doi:10.13026/3z8q-x658), Nutrition5k (CC BY 4.0) and USDA FoodData Central SR Legacy (public domain). The data splits, the reference values of the 100 annotated meals, the prompts, the value of every reported result, the analysis code and the LoRA adapter are available from the corresponding author on reasonable request. Results can be browsed read-only at https://glycorepas-dt2.vercel.app (username: lecteur; password: repas-texte-glucides).

https://figshare.com/articles/dataset/21600933

https://physionet.org/content/cgmacros/1.0.0/

https://github.com/google-research-datasets/Nutrition5k

https://fdc.nal.usda.gov/download-datasets

## Declarations

### Ethics statement

This secondary analysis used only publicly available, de-identified data released by their original investigators (ShanghaiT2DM [22]; CGMacros [25]) and food composition data that involve no human participants (Nutrition5k [24]; USDA FoodData Central [27]). The approvals and consents of the original data collections are described in the corresponding data descriptors. No participant was recruited, no new data were collected and no attempt was made to re-identify individuals. Ethical approval was therefore not required for the present analysis.

### Author contributions (CRediT)

MNBA (lead author): conceptualisation, methodology, investigation, data curation, software, formal analysis, validation, visualisation, project administration, writing – original draft, writing – review and editing. NL, BBA, SN and MAA: clinical interpretation, writing – review and editing. All authors approved the submitted version.

### Data availability

All data used are publicly available: ShanghaiT2DM (figshare, CC BY 4.0) [22], CGMacros (PhysioNet) [25], Nutrition5k (CC BY 4.0) [24] and USDA FoodData Central SR Legacy (public domain) [27]. The frozen data splits, the reference values of the 100 annotated meals, the prompts, the value of every reported result and the LoRA adapter are available from the corresponding author on reasonable request. The results can be browsed, read-only, on the online dashboard https://glycorepas-dt2.vercel.app (username: lecteur; password: repas-texte-glucides).

### Code availability

The analysis code (Python), the Kaggle training and inference notebooks and the LoRA adapter produced by QLoRA (Apache 2.0 licence) are available from the corresponding author on reasonable request.

### Protocol and registration

The protocol (definitions, splits, outcomes, analyses and the step-1 pass criterion) was written and the splits frozen before training (section 2.1), but it was not deposited with a public time stamp. The study was not registered in a public registry. The status of each analysis (main, secondary, exploratory or sensitivity) is given in Table S1.

### Patient and public involvement

Patients and the public were not involved in the design, conduct, reporting or dissemination plans of this research.

### Software tools

The analysis was entirely programmed in Python; a programming assistant (Claude, Anthropic) helped write this code and format the manuscript. The lead author reviewed the code and checked the correspondence tables and every reference value, and the authors take full responsibility for the content.

## Acknowledgements

We thank the investigators who released the ShanghaiT2DM, CGMacros and Nutrition5k datasets, and Kaggle for access to GPUs.

## Supplementary material

**Table S1.**
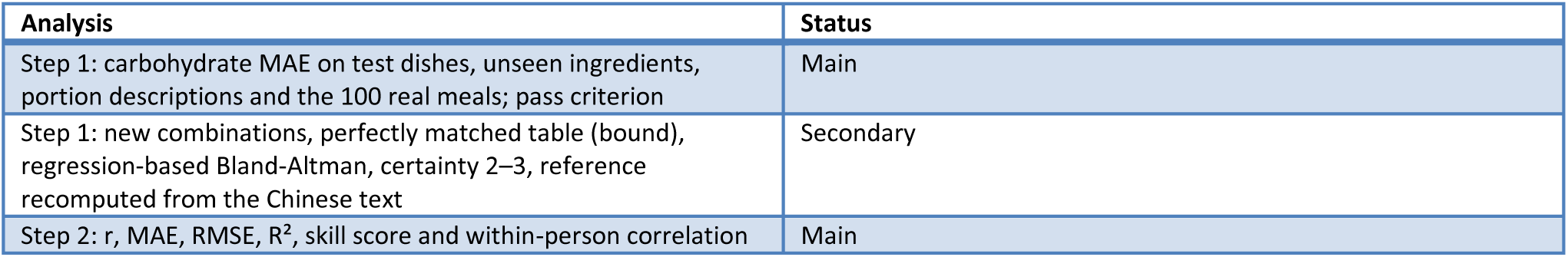
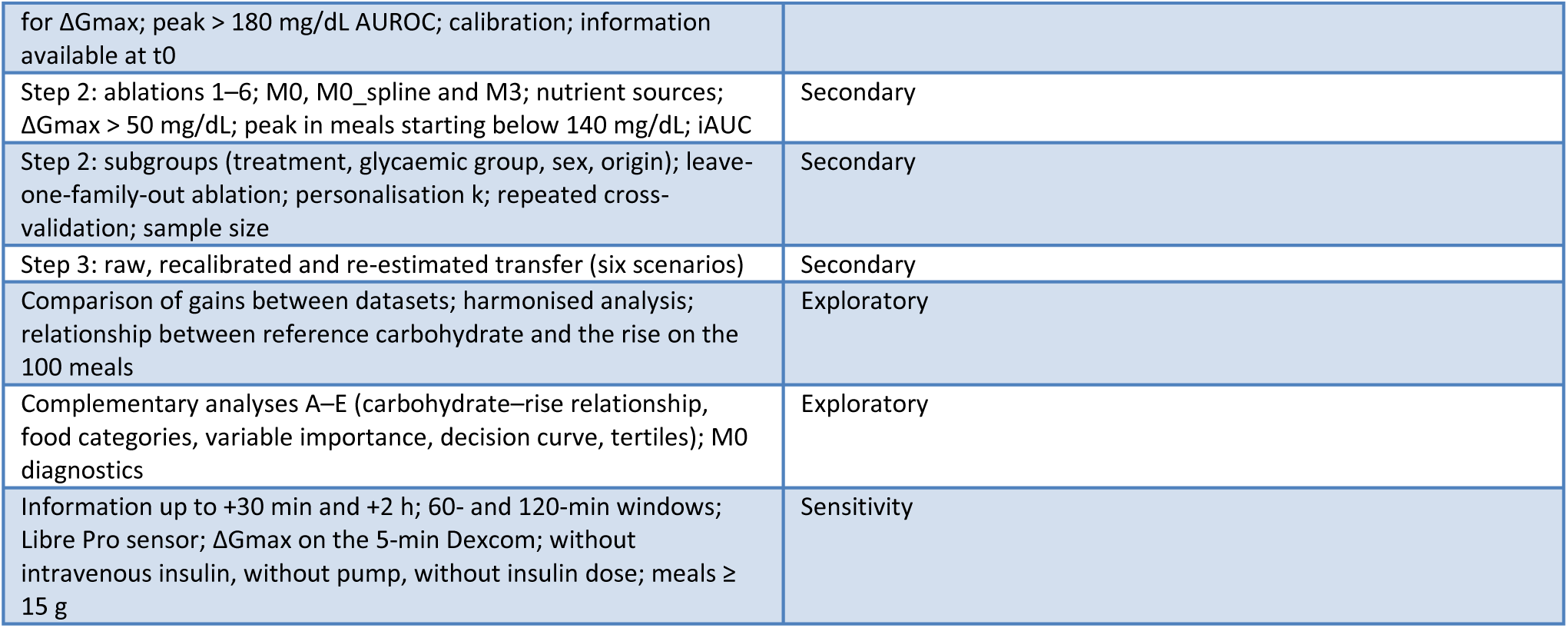
Status of the analyses.

**Table S2.** The main analysis (strict t0) and the sensitivity analysis (information up to +30 min and +2 h) are compared, model by model, in the file resultats/tables/t0_vs_actuel.csv of the repository.

### LLM prompt (nutriments_v1, frozen)

System: “You are a clinical dietitian. You estimate the nutrient content of a meal from its written description, using standard food composition tables (USDA FoodData Central). Quantities may be given in grams, millilitres or household portions. Reply with one JSON object only, no text before or after, with exactly these keys (numbers, no units): glucides_g, proteines_g, lipides_g, kcal, masse_g.” User: “Meal: {texte}”. Repair: “Your previous answer could not be used ({raison}). Reply again with one JSON object only, with exactly the keys glucides_g, proteines_g, lipides_g, kcal, masse_g and numeric values >= 0.”

### iAUC computation

At each grid time, glucose minus baseline is set to 0 if negative, then the area is computed by the trapezoidal rule from t0; segments below baseline are not subtracted and crossing points with baseline are not interpolated.

### LoRA adapter hash (SHA-256)

b2bca730bb1e3bc7f27a0ef32b0801feb3b1b772c151bd68d153a48e85bf6f28 (base model: Qwen3-4B, main branch on Hugging Face at training time).

**Figure S1.**
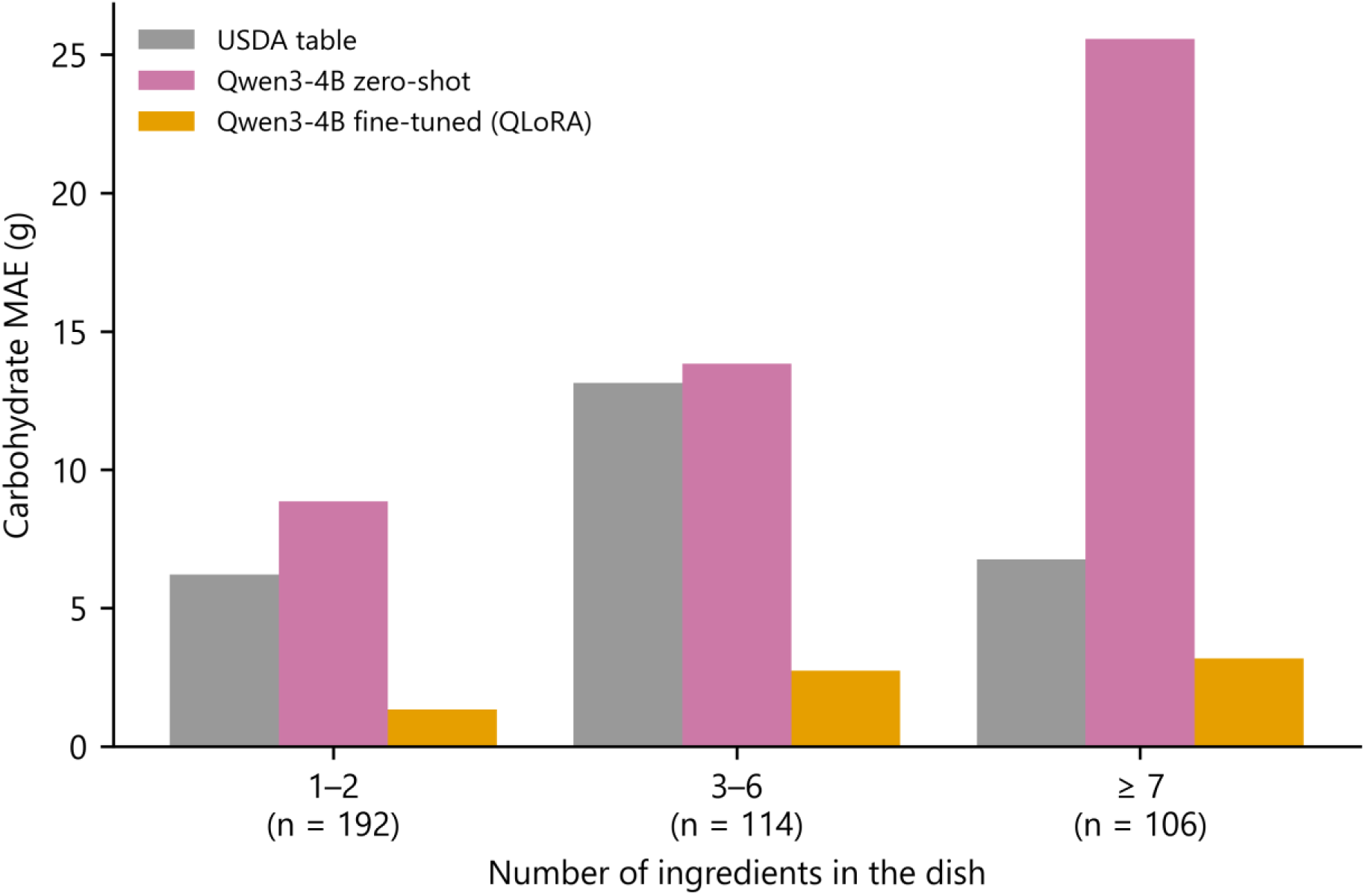
Carbohydrate MAE by number of ingredients in the dish (Nutrition5k, test; counts per class). The automatically matched USDA table is not monotonic: its error depends mainly on the quality of name matching.

**Figure S2.**
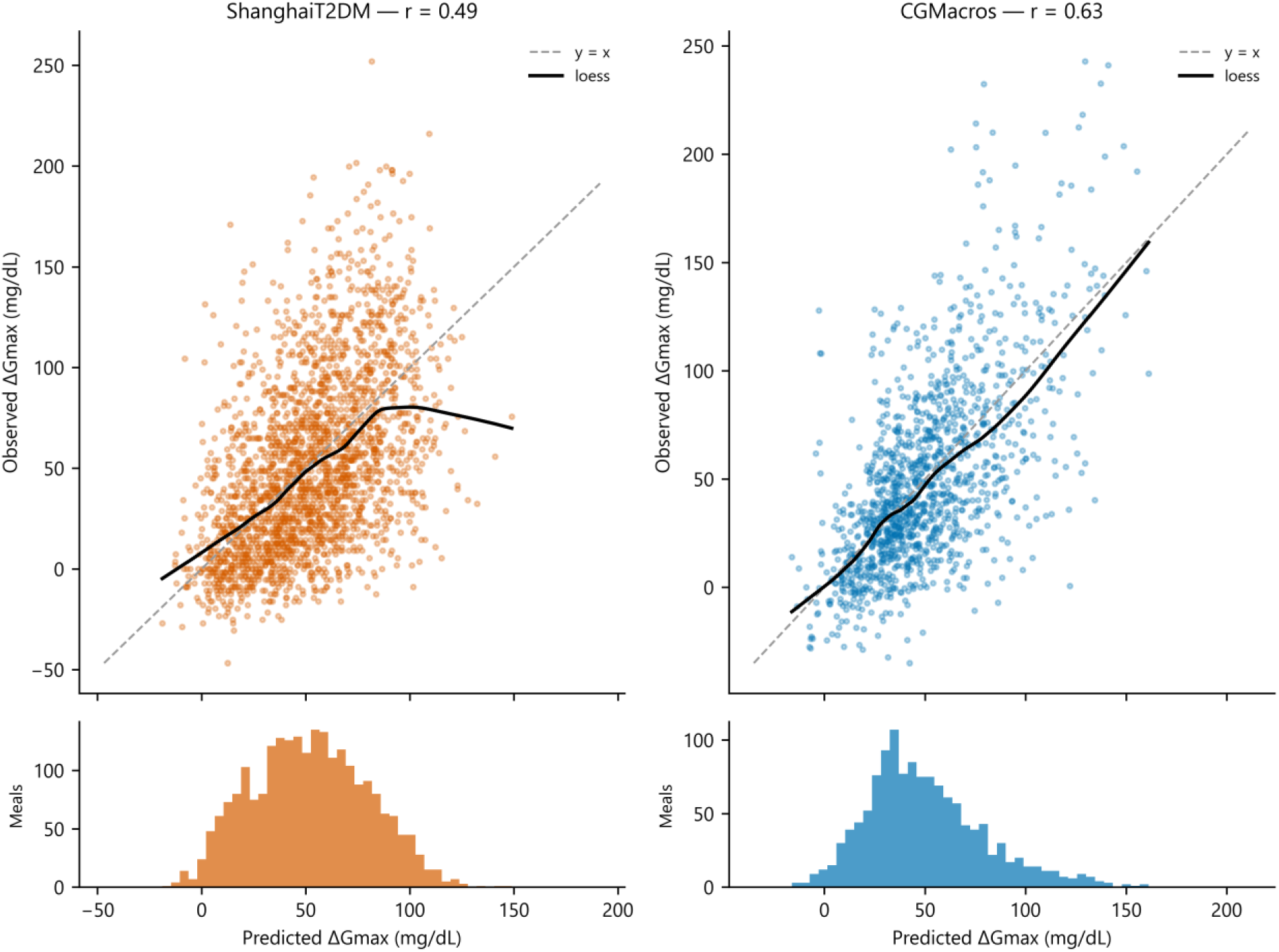
Observed versus M2-predicted ΔGmax (y = x line and loess smoothing), with the distribution of predictions.

**Figure S3.**
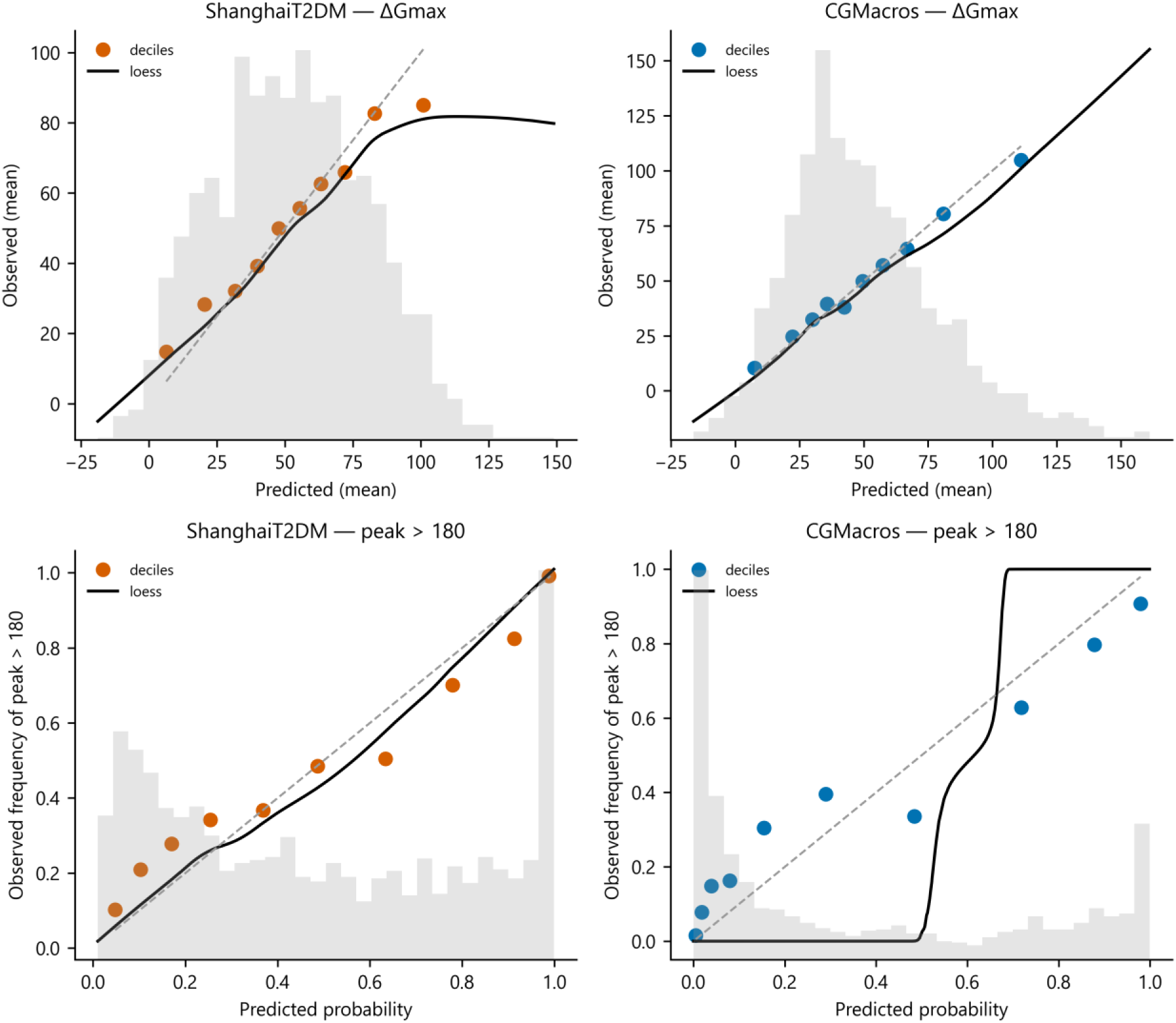
Calibration of M2 (ΔGmax and peak > 180 mg/dL): prediction deciles, loess smoothing and distribution of predictions.

**Figure S4.**
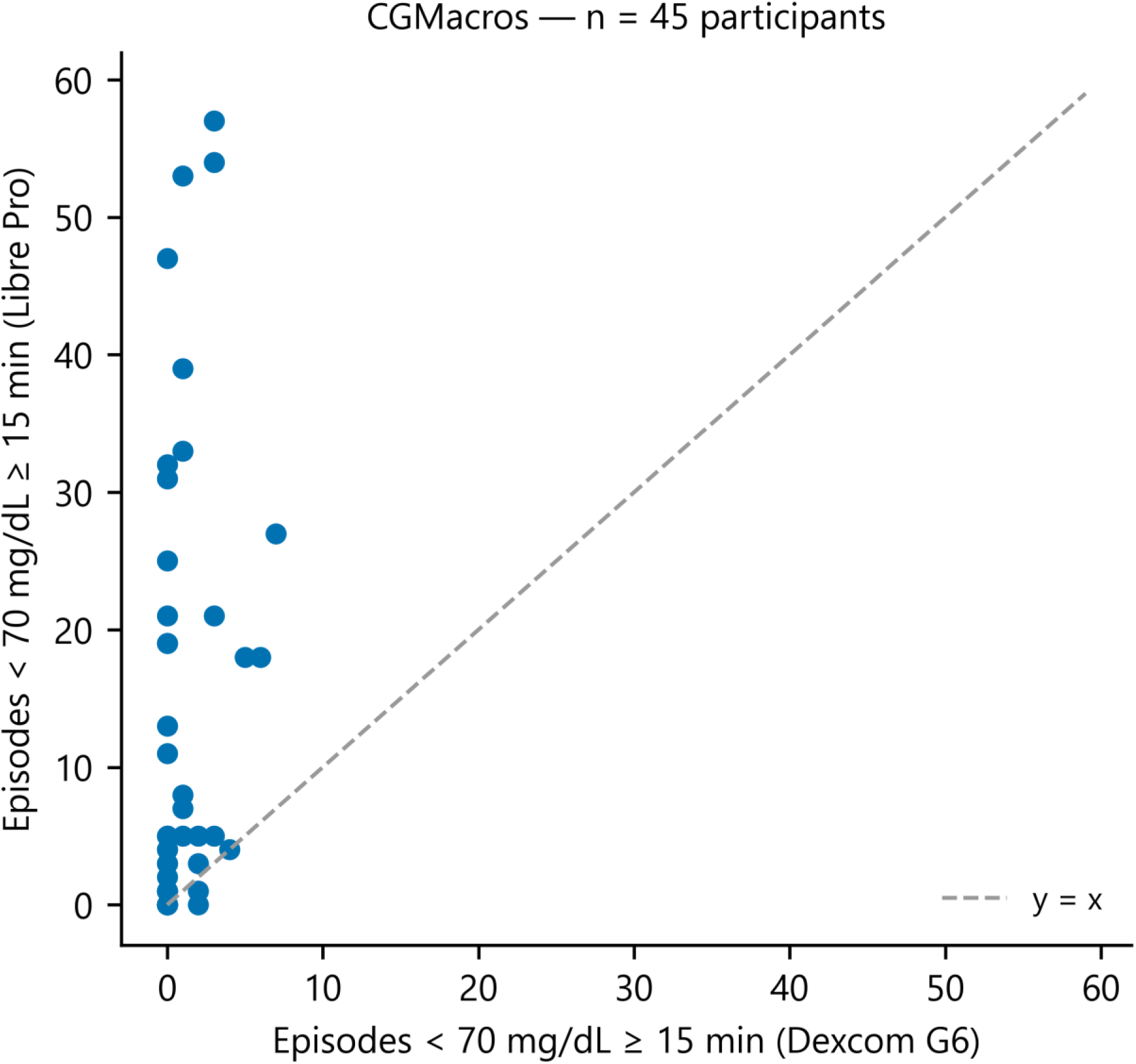
Episodes < 70 mg/dL seen by the Libre Pro and by the Dexcom G6 in the same CGMacros people (y = x line).

**Figure S5.**
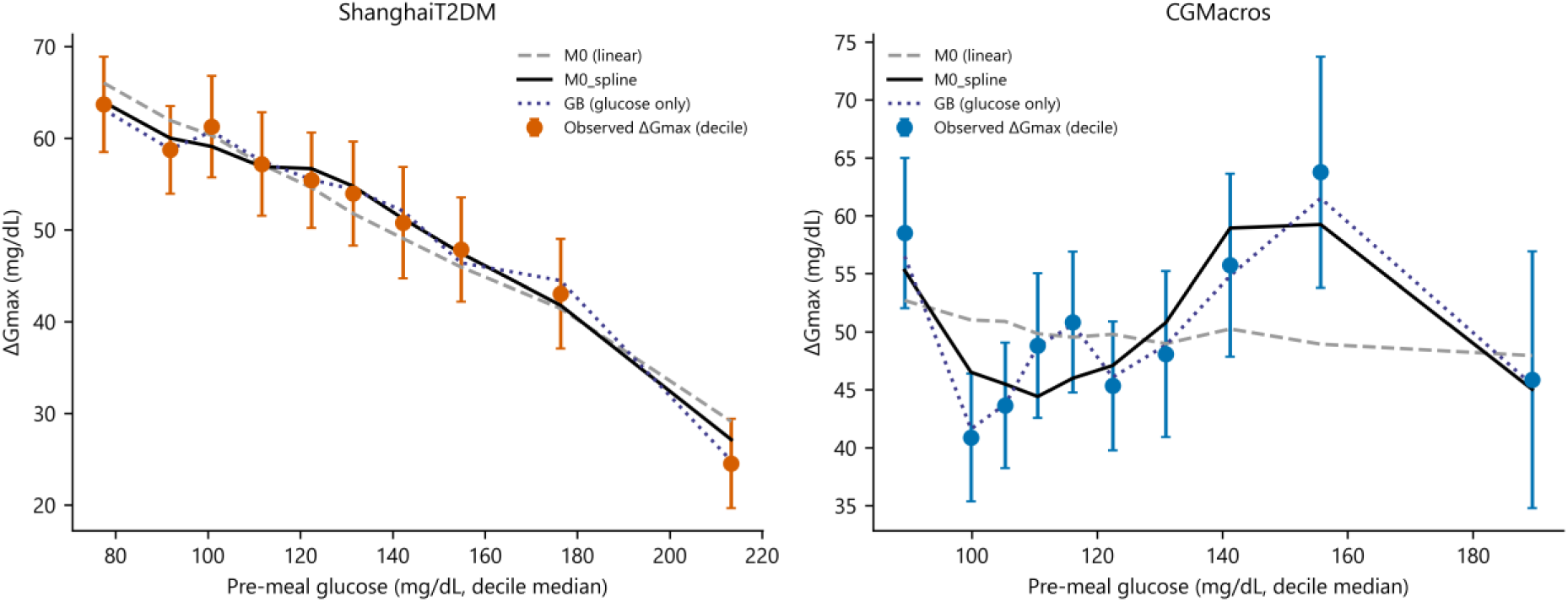
Mean ΔGmax by decile of starting glucose (95% CI) and predictions of M0, M0_spline and gradient boosting with glucose alone.

**Figure S6.**
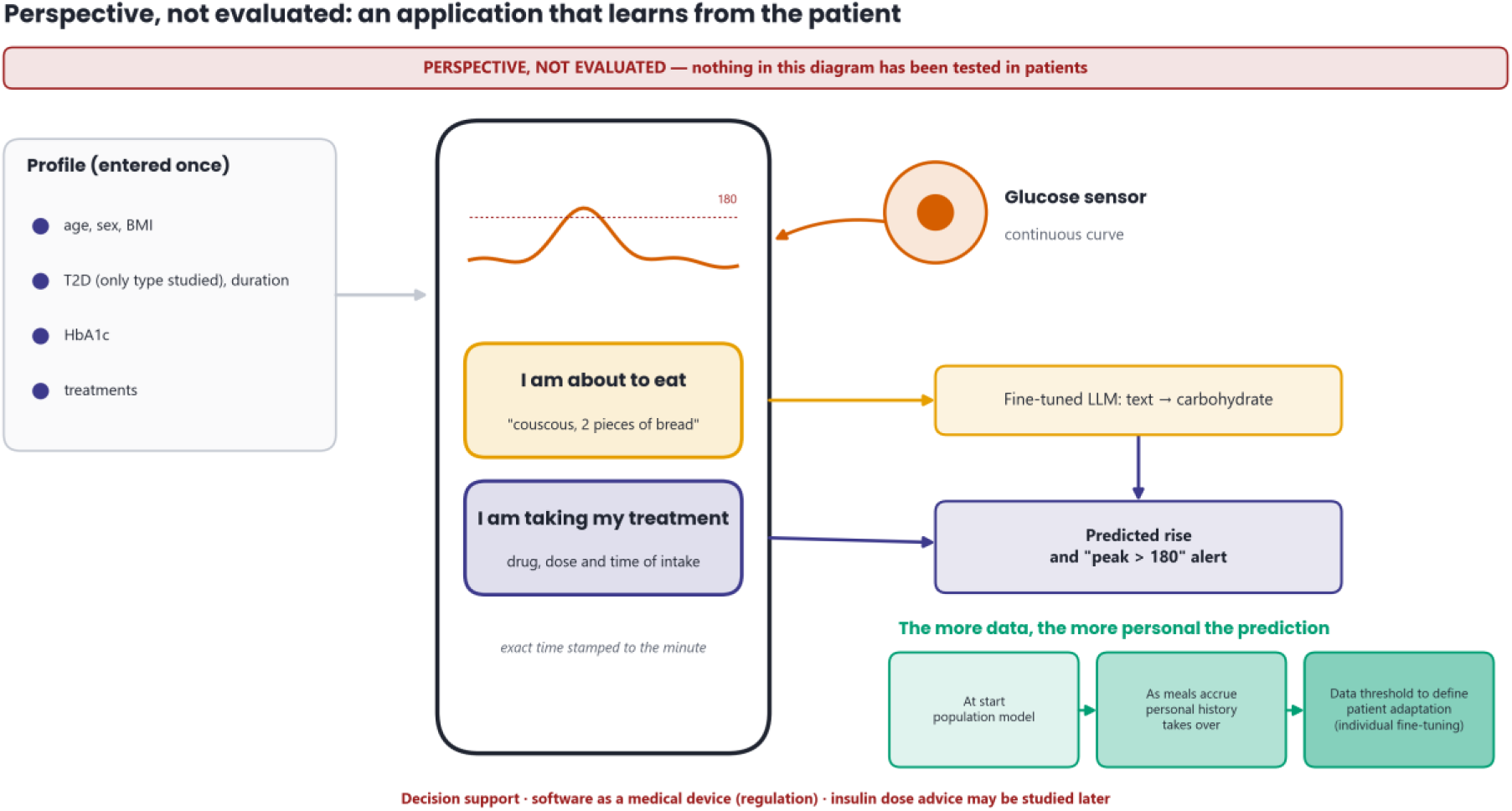
Perspective, not evaluated: monitoring application coupled with a glucose sensor (decision aid).

## Online dashboard

The data, results and cases of this study can be browsed on an online dashboard, GlycoRepas: patient meals, Nutrition5k dishes, the 100 annotated meals, the results of the three steps and an interactive map of the study. Access is read-only: no file can be downloaded from it.

- Address: https://glycorepas-dt2.vercel.app
- Username: lecteur
- Password: repas-texte-glucides

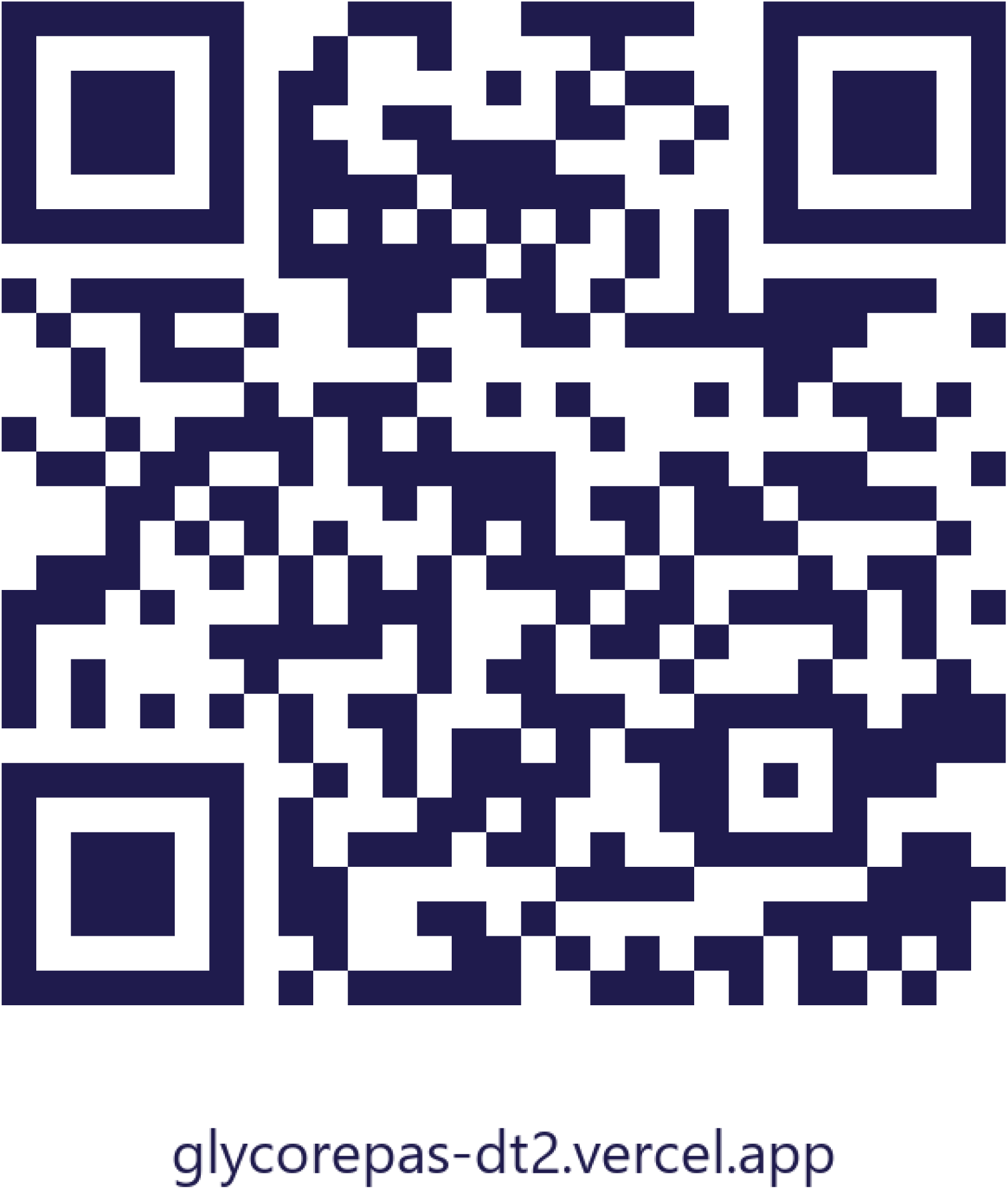

*QR code giving access to the GlycoRepas dashboard*.

## Summary sheets

The following four sheets summarise the study, each on one page: the datasets and their inclusion and exclusion criteria, the step-1 models and the sets held out from training, the step-2 configurations and the transfer, then the key messages.

**Summary sheet 1.**
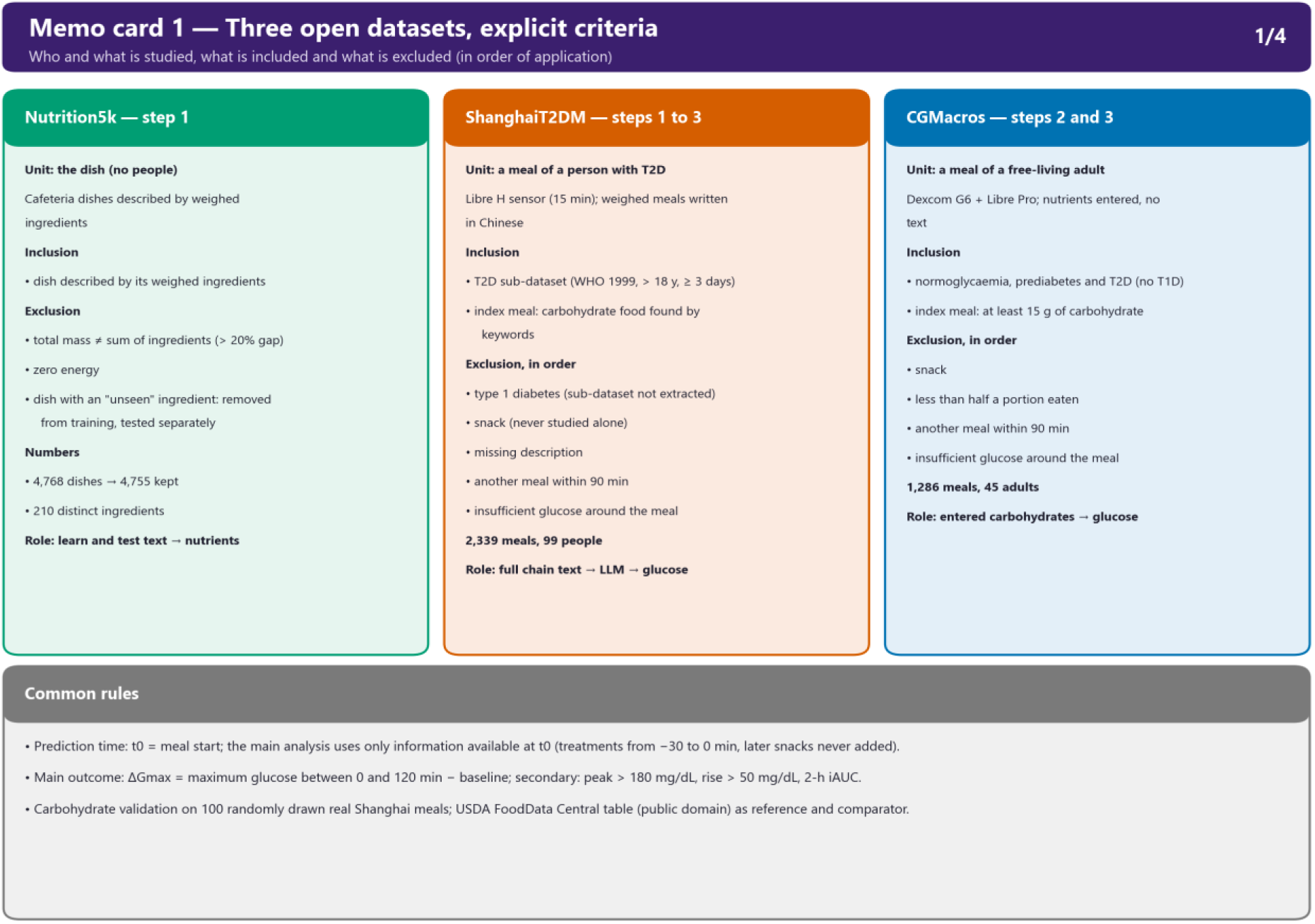
Three open datasets: unit of analysis, inclusion and exclusion criteria in order of application, numbers and role of each dataset; common rules.

**Summary sheet 2.**
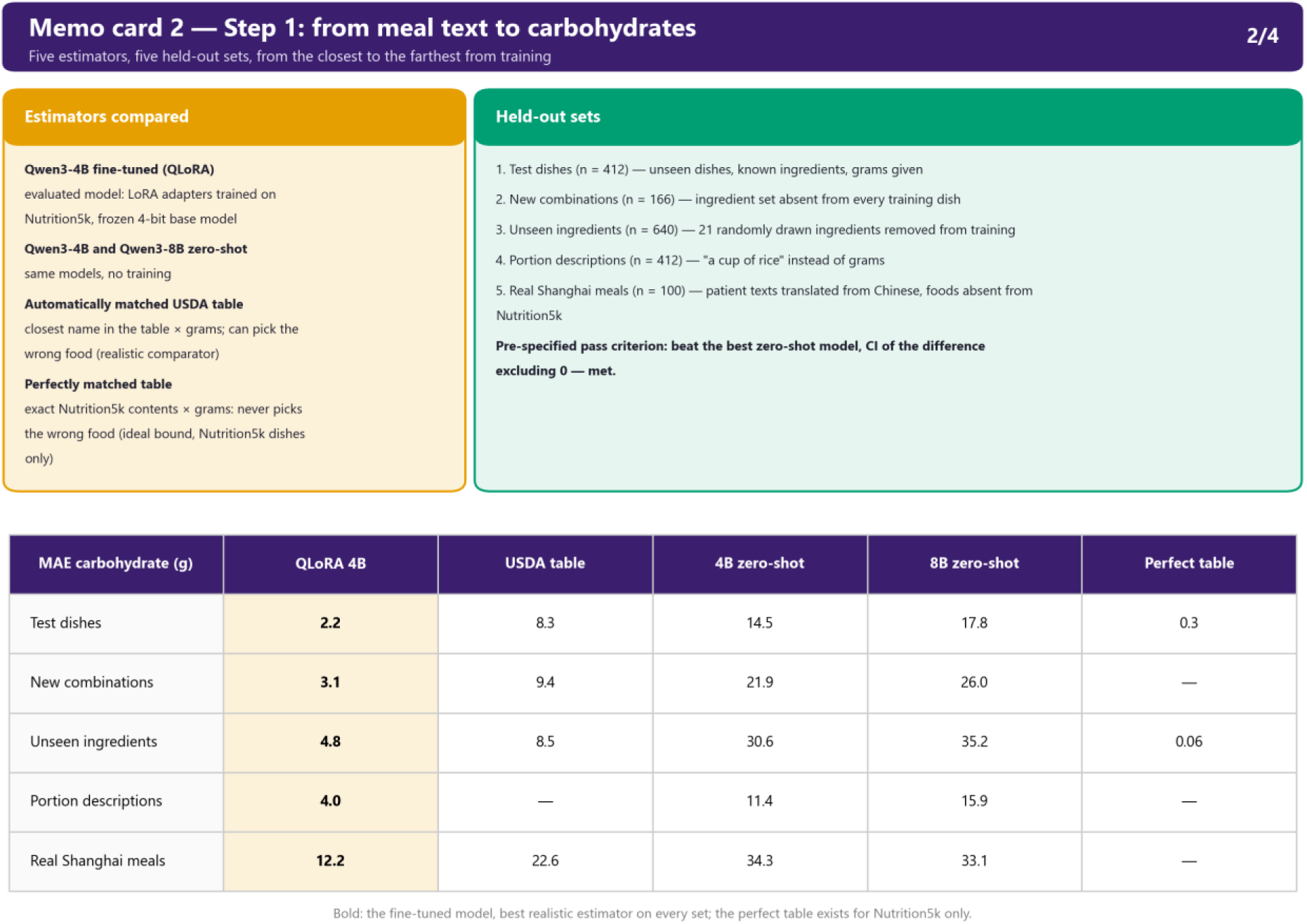
Step 1: compared estimators, sets held out from training (from closest to furthest) and carbohydrate MAE of each estimator on each set.

**Summary sheet 3.**
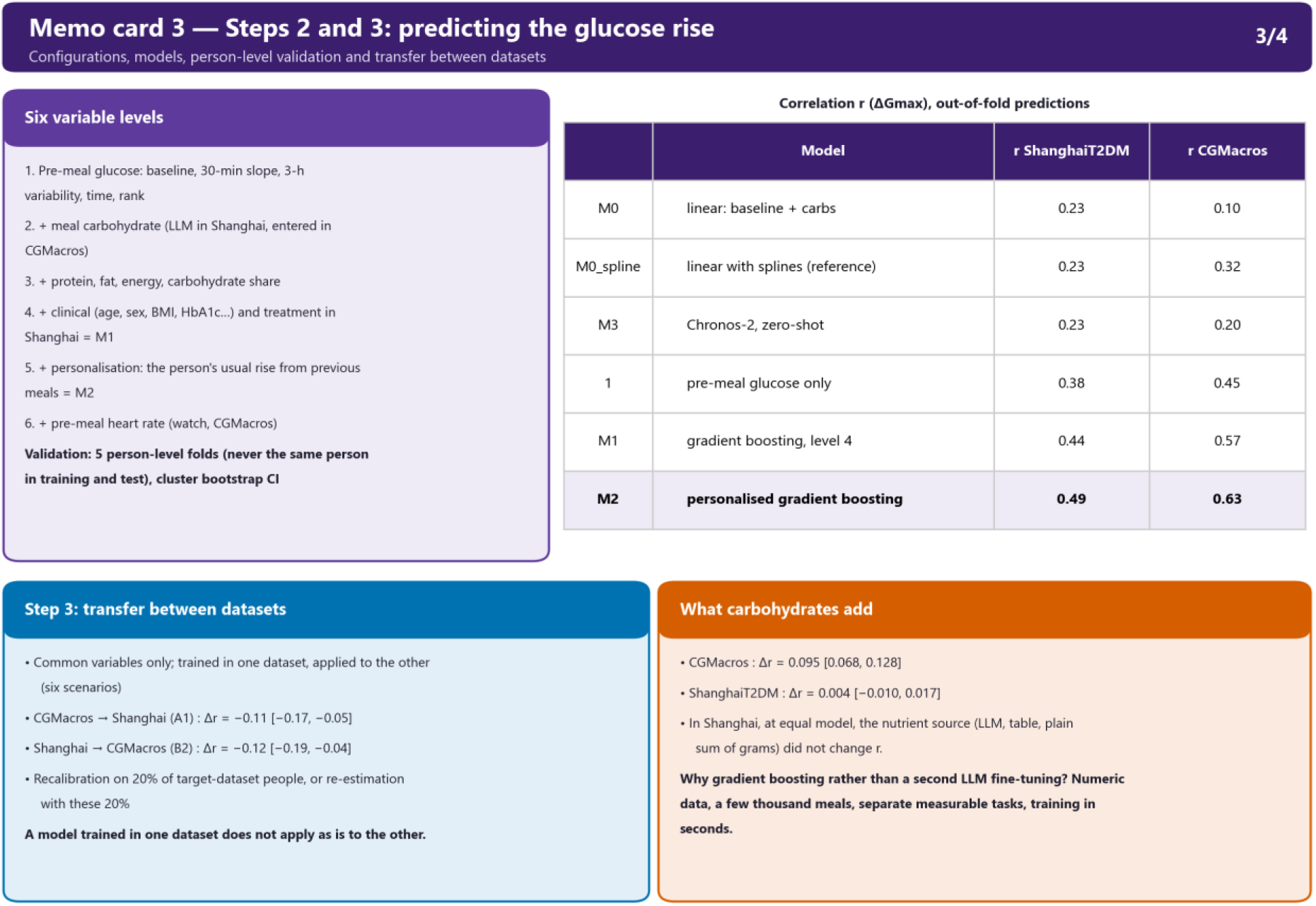
Steps 2 and 3: six variable levels, models M0 to M3 and their out-of-fold correlation, contribution of carbohydrate and transfer between datasets.

**Summary sheet 4.**
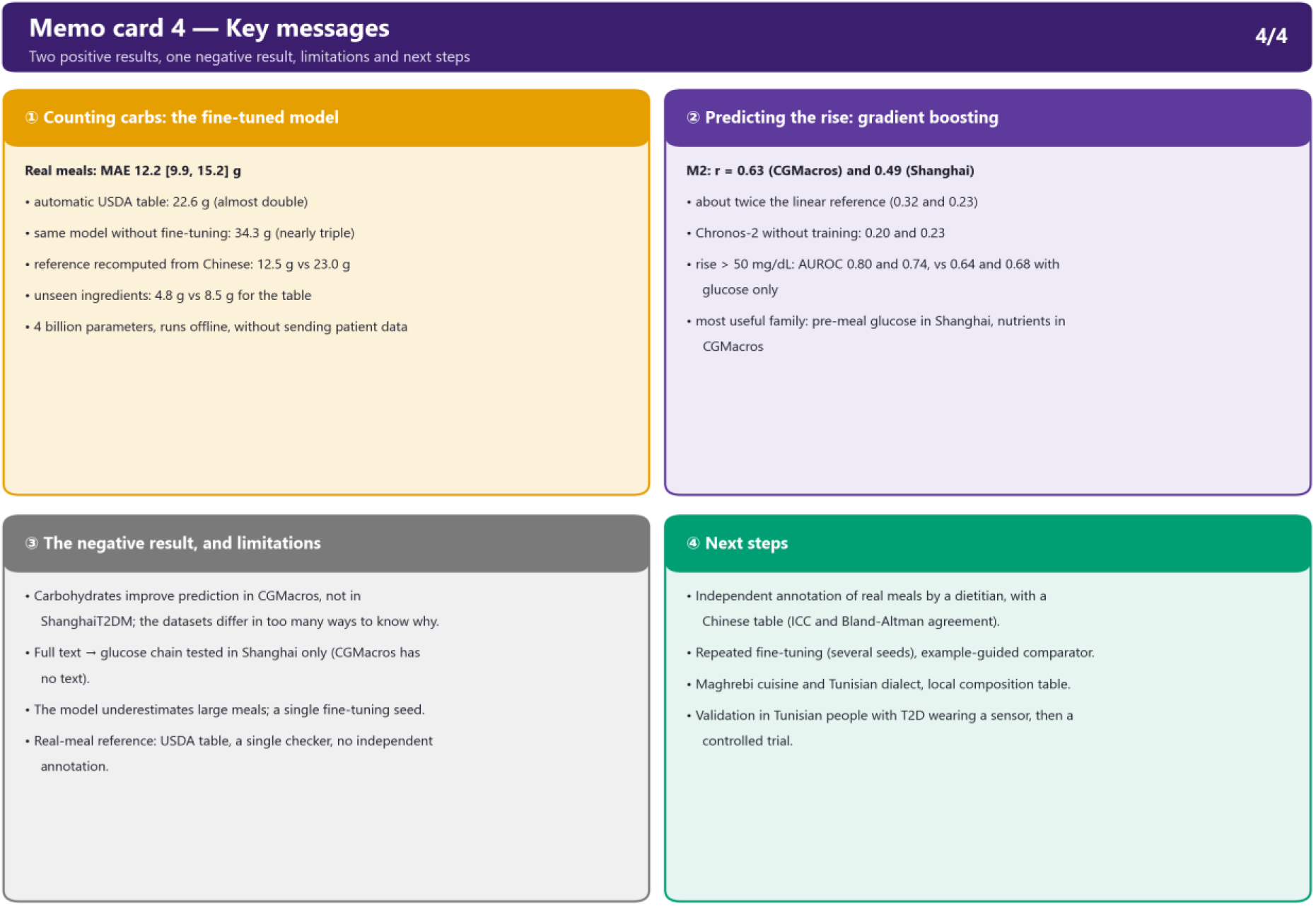
Key messages: the two positive results, the negative result, the limitations and the next steps.

