## Supplementary_TRIPOD-AI_checklist for "From meal text to carbohydrate to postprandial glucose rise: QLoRA fine-tuning of the Qwen3-4B language model and gradient-boosting prediction in type 2 diabetes"

### TRIPOD+AI checklist: where each item is reported

*Completed by the authors. Item numbers follow the TRIPOD+AI checklist (Collins et al., BMJ 2024;385:e078378); item labels are abbreviated. Section, table and figure numbers refer to the full manuscript (preprint).*

| Item | Topic | Where reported |
|---|---|---|
| **1** | Title | Title page (title and subtitle: development and validation study, target population, outcome) |
| **2** | Abstract | Abstract |
| **3a** | Background: health-care context and rationale | 1. Introduction |
| **3b** | Target population and intended purpose | 1. Introduction (objectives); 4.7 |
| **3c** | Known health inequalities | 2.6 (subgroups by sex and origin); 3.3 (Subgroups); 4.5 (limitation 7) |
| **4** | Objectives | 1. Introduction, last paragraph |
| **5a** | Data sources | 2.2; Table 1 |
| **5b** | Dates of data collection | 2.1 (recruitment periods of ShanghaiT2DM and CGMacros; recording dates of ShanghaiT2DM) |
| **6a** | Setting | 2.1; 2.2 |
| **6b** | Eligibility criteria | 2.2; Table 2 |
| **6c** | Treatments received | 2.6 (treatment predictors); Table 5 |
| **7** | Data preparation and quality checking | 2.2 (merging of intakes, snacks); 2.3 (15-min grid); 2.5 (splits, augmentation, JSON schema validation); 2.1 (SHA-256) |
| **8a** | Outcome definition | 2.3 (ΔGmax, iAUC, peak > 180 mg/dL, rise > 50 mg/dL); 2.5 (reference carbohydrate of real meals) |
| **8b** | Subjective outcome assessment | 2.5 (reference of the 100 real meals: certainty levels, line-by-line checking) |
| **8c** | Blinding of outcome assessment | 2.5 (reference checked without access to LLM outputs) |
| **9a** | Choice of predictors | 2.6; Table 4 |
| **9b** | Definition of predictors | 2.6 |
| **9c** | Subjective predictor assessment | 2.2 and 2.6 (nutrients estimated by the LLM in Shanghai, recorded estimates in CGMacros); 4.5 (limitations) |
| **10** | Sample size | 2.6 (Sample size: set by the public datasets, adequacy checked with the Riley et al. method); 3.3; 4.5 (limitation 7) |
| **11** | Missing data | 2.6 (Missing data: native handling by gradient boosting, median imputation learned on training folds); 3.1 (frequency of missing values); 2.5 (invalid LLM output) |
| **12a** | Use of data for development and evaluation | 2.1; 2.5 (dish-level splits); 2.6 (person-level 5-fold cross-validation); 2.7 (transfer) |
| **12b** | Handling of predictors | 2.6 |
| **12c** | Model type, building steps, hyperparameters, internal validation | 2.4 (technical details of both models); 2.5 (QLoRA hyperparameters); 2.6 (nested cross-validation) |
| **12d** | Heterogeneity across clusters | 2.6 (subgroups, between-dataset comparison); 2.10 (cluster bootstrap) |
| **12e** | Performance measures | 2.5 (step-1 measures); 2.6 (r, MAE, calibration, AUROC, Brier score, decision curve); Box 1 |
| **12f** | Model updating | 2.7 (recalibration and re-estimation on 20% of the target |

| | | |
|---|---|---|
| | | dataset) |
| **12g** | Calculation of predictions for evaluation | 2.6 (out-of-fold predictions) |
| **13** | Class imbalance | Primary outcome continuous; peak frequency reported in 3.1 |
| **14** | Fairness | 2.6 and 3.3 (performance by glycaemic group, treatment class, sex and origin) |
| **15** | Model output | 2.4 and 2.6 (predicted ΔGmax in mg/dL; probability of peak > 180 mg/dL) |
| **16** | Differences between development and evaluation data | 2.2 (Table 1); 2.7; 3.4 |
| **17** | Ethical approval | 2.1 (Ethics); Declarations (Ethics statement) |
| **18a** | Funding | Declarations |
| **18b** | Conflicts of interest | Declarations |
| **18c** | Protocol | 2.1; Declarations |
| **18d** | Registration | Declarations (not registered) |
| **18e** | Data sharing | Declarations (Data availability; online dashboard) |
| **18f** | Code sharing | Declarations (Code availability) |
| **19** | Patient and public involvement | Declarations |
| **20a** | Flow of participants | 3.1; Figure 2 |
| **20b** | Characteristics of participants | 3.1; Table 5 |
| **20c** | Comparison of predictor distributions between datasets | Table 5 (Shanghai versus CGMacros, relevant to transfer) |
| **21** | Participants and outcome events per analysis | 3.1; Tables 5 to 7 |
| **22** | Full model specification | 2.4 to 2.6; adapter hash in the supplementary material; code available (Declarations) |
| **23a** | Model performance with CIs, including subgroups | 3.2; 3.3; Tables 6 and 7 |
| **23b** | Heterogeneity of performance across clusters | 3.3 (Subgroups); 3.4 (transfer) |
| **24** | Results of model updating | 3.4 |
| **25** | Interpretation | 4.1 to 4.4 |
| **26** | Limitations | 4.5 |
| **27a** | Handling of poor-quality or unavailable input data | 2.5 (schema validation and second attempt); 2.6 (missing data) |
| **27b** | User interaction with input data | 4.7 (meal described in words by the patient) |
| **27c** | Next steps for future research | 4.7 |
